# Modelling upper respiratory viral load dynamics of SARS-CoV-2

**DOI:** 10.1101/2021.05.01.21256182

**Authors:** Joseph D. Challenger, Cher Y. Foo, Yue Wu, Ada W. C. Yan, Mahdi Moradi Marjaneh, Felicity Liew, Ryan S. Thwaites, Lucy C. Okell, Aubrey J. Cunnington

## Abstract

Relationships between viral load, severity of illness, and transmissibility of virus, are fundamental to understanding pathogenesis and devising better therapeutic and prevention strategies for COVID-19. Here we present within-host modelling of viral load dynamics observed in the upper respiratory tract (URT), drawing upon 2172 serial measurements from 605 subjects, collected from 17 different studies. We developed a mechanistic model to describe viral load dynamics and host response, and contrast this with simpler mixed-effects regression analysis of peak viral load and its subsequent decline. We observed wide variation in URT viral load between individuals, over 5 orders of magnitude, at any given point in time since symptom onset. This variation was not explained by age, sex, or severity of illness, and these variables were not associated with the modelled early or late phases of immune-mediated control of viral load. We explored the application of the mechanistic model to identify measured immune responses associated with control of viral load. Neutralizing antibody correlated strongly with modelled immune-mediated control of viral load amongst subjects who produced neutralizing antibody. Our models can be used to identify host and viral factors which control URT viral load dynamics, informing future treatment and transmission blocking interventions.

## Introduction

COVID-19 exhibits a wide range of severity, from asymptomatic infection to severe disease leading to hospitalisation and death. Age and sex have emerged as important risk factors for poor outcome (1,2). Viral load in the respiratory tract has been reported as an additional determinant of severity of illness (3,4) and also a determinant of likelihood of transmission (5). However, viral load varies over the course of illness due to dynamic interaction with the host immune response, and measurements at single points in time provide limited insight into this dynamic process. Within-host models of viral load can help to distinguish the sequence of events by tracking both viral dynamics and host response over time, accounting for the effect of multiple factors simultaneously (4,6,7).

Studies measuring viral load over time in COVID-19 are beginning to establish viral dynamics and explore correlates of protection in the host response, although findings to-date are somewhat contradictory and these relationships are still not well characterised (8). Viral load in the upper respiratory tract (URT) peaks early in infection, usually before or within a few days of symptom onset (8–12). Some studies suggest that viral load in the lower respiratory tract (LRT) may peak later, in the second week after symptoms (9), but this is much harder to measure in a serial manner. Viral load at a given time after diagnosis or detection tends to be similar between asymptomatic and symptomatic cases (13), but evidence tends towards a longer duration of viral shedding in more severe cases and older individuals (14,15). Despite detection of viral material in samples from some individuals several weeks after onset of symptoms, infectious virus is not usually present beyond 8-14 days (16–19).

The host response during COVID-19 has many features typical of an anti-viral immune response, including the generation of antibodies and T cell populations (20). Antibodies are chiefly considered to contribute to viral clearance through pathogen neutralisation, though other effector functions may be of significance to COVID-19 (21). A diverse array of T cell populations are active during COVID-19, including cytotoxic and helper populations. While these cells contribute to viral clearance, emerging data indicates that T cells may also contribute to immunopathology in severe cases of COVID-19 (22). Additionally, COVID-19 is associated with lymphopenia in peripheral blood (23), possibly reflecting migration of cells to the site(s) of viral infection. In addition to these adaptive immune processes, innate immune processes are considered to play a major part in the pro-inflammatory state that scales with COVID-19 severity (24).

Within-host models are being developed to characterise viral kinetics in relation to host responses and disease outcomes and to guide therapeutic development. For example, Néant et al. (25) found an association between higher viral load late in infection and mortality. Goyal et al. (26) inferred different stages of host response from observing three stages of viral decline: a rapid drop following peak viral load, a period of slower decline, then rapid elimination of the virus. Benefield et al. estimated that viral load peaks prior to symptoms, suggesting substantial pre-symptomatic transmission (27). Other within host models have been used to explore the potential effects of antivirals, immunotherapies, and prophylactic treatment (28,29). More detailed models have simulated viral load in different tissues and detailed components of the innate and cellular immune response (30,31).

Many modelling studies to date have been calibrated to limited longitudinal data on viral load and particularly on host responses, which reduces parameter identifiability and the ability to infer pathways of pathogenesis and protection. Here, we first used linear regression models to assess the associations of age, sex, and severity of disease with viral load. Then we developed a model of viral dynamics, in which we pragmatically represented the complex host response to the virus in two phases: an early phase which restricts the initial rate of viral replication, and a later phase which acts to clear virus. Fitting this model to longitudinal viral load measurements from previously published studies allowed us to make individualised estimates of key metrics such as peak viral load and rate of decline in viral load after the peak, which could also be related to age, sex, and severity. This model represents the first step towards a well-validated, flexible, and open-source framework which can be utilised to better interpret immune responses in COVID-19 in the context of viral load, and to understand how different treatments given at different stages of illness might influence viral load, transmission potential, host response, and outcome. We illustrate this with an example immune response dataset.

## Results

Our literature search revealed 53 studies reporting longitudinal viral load measurements. We successfully obtained individual-level viral load measurements from 19 studies (either from the supplementary materials of the publication or preprint, or by emailing the corresponding authors). The analysis presented here utilises data from 17 of these studies (summarised in Table 1). Studies that were identified but did not provide data are shown in Supplementary Table 1. We excluded two studies that contained little or no longitudinal data for URT samples (32,33). In all studies, a description of disease severity for each patient was available, although the level of detail varied between studies. In six studies viral loads were fully quantified from concurrent standard curves, whilst in the remaining 11 studies cycle-threshold (Ct) values were reported. To combine data from different studies we generated an “average” standard curve, using data from 7 previously reported standard curves to convert Ct values to viral load per ml (Supplementary Figure 1). Details on antiviral or immunomodulatory treatments were available for some studies (Supplementary Table 2).

**Table 1.**
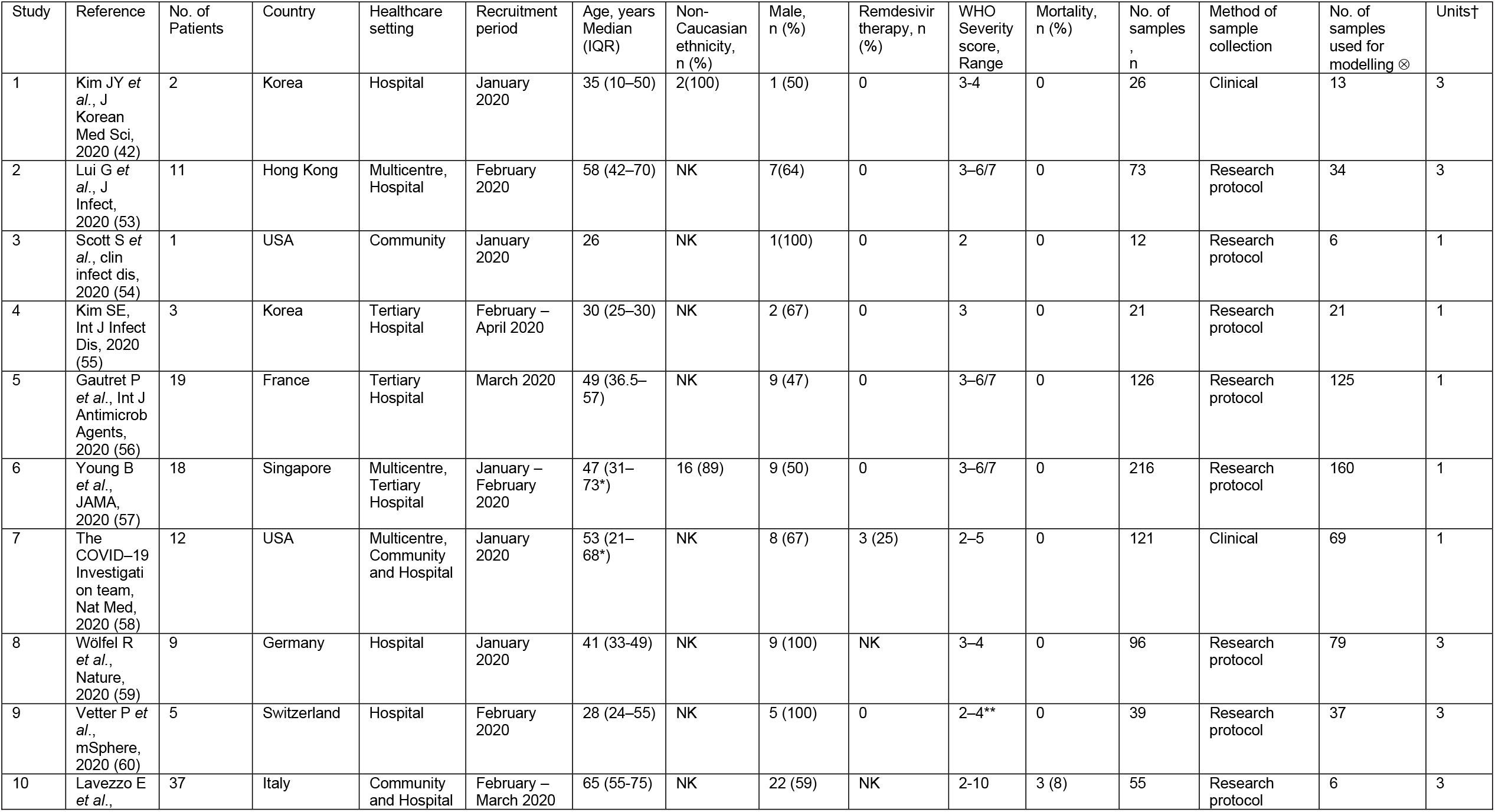
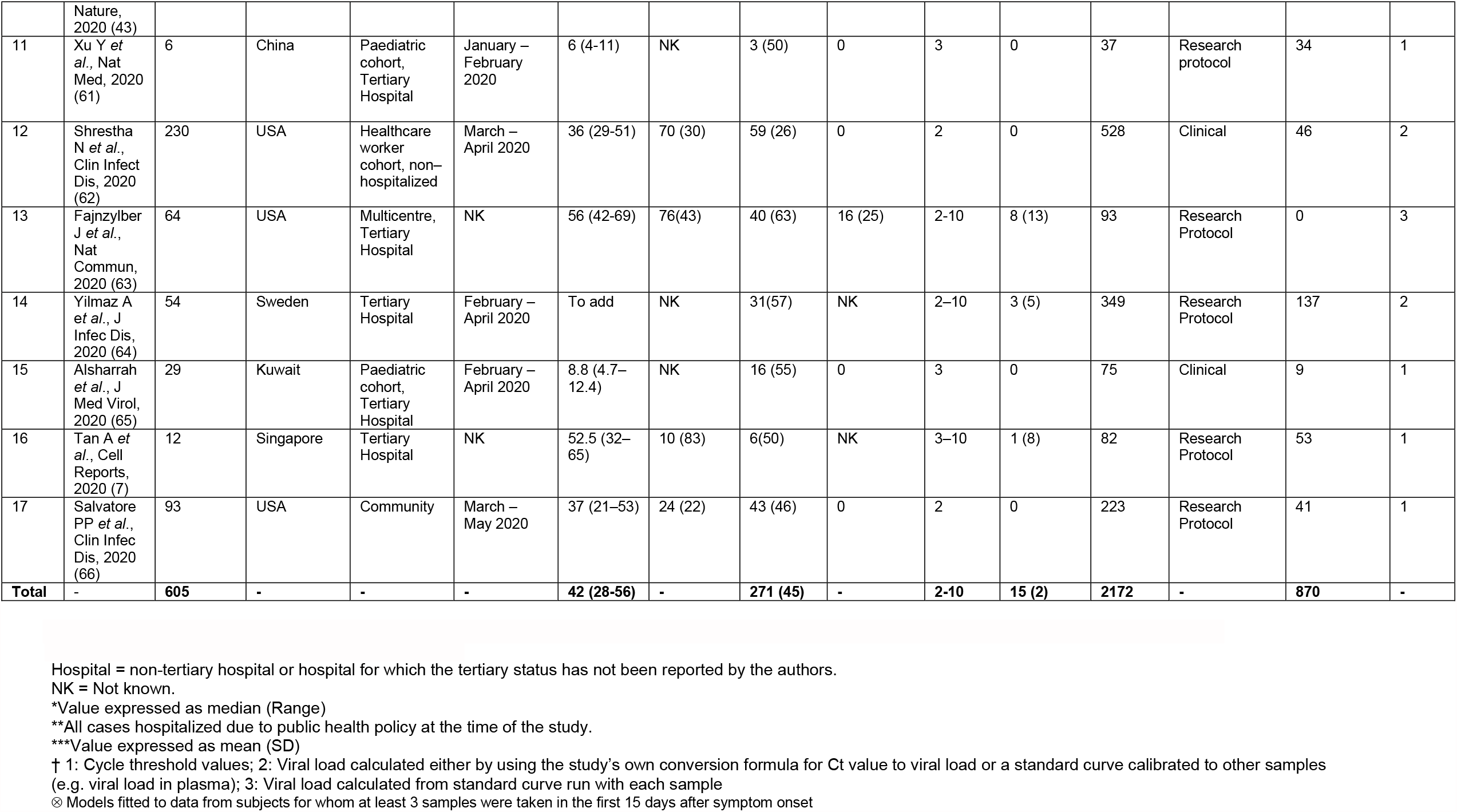
Summary of participant characteristics and sample collection protocols for each study, from which data was collected to model viral load dynamics. Only cases of symptomatic COVID-19 were included.

In total, 2172 URT samples from 605 patients were used for analysis. Subject-level data on age and sex were available for 576 subjects and missing for 29 subjects. The majority (492 out of 576; 85%) of subjects were under 60, compared to 84 (15%) aged 60 or over; 321 (55%) patients were female. Disease severity was categorised using the WHO scale (34), where 501 (83%) patients (contributing 1698 (78%) samples) experienced mild COVID-19 illness, 65 (11%) patients (contributing 359 (17%) samples) had moderate severity illness, and 39 (6%) patients (contributing 115 (5%) samples) had severe illness. Supplementary Figure 2 shows the viral load data for all patients, presented separately for each study. The vast majority (2163, or 99.6%) of samples were taken after the onset of symptoms, although samples were collected from a minority (8 patients across all 17 studies) prior to symptom onset e.g. if they were identified as a close contact of another patient. The median timing of swabs was 12 days after symptom onset (interquartile range: 6-19; range: 2 days before symptom onset to 54 days afterwards). Pooling the data from all studies, we found that the median viral load peaked one day after symptom onset (Figure 1a), although less data was available on the day of symptom onset compared to subsequent days (Figure 1c). 421 patients had more than one URT swab recorded: for 60.1% of these patients, the first sample had the highest recorded viral load. Viral load estimates at corresponding times after onset of symptoms did not differ systematically between studies in which viral load was calculated by the authors of the original study or inferred from our averaged standard curve (Figure 1b), although less data was contributed by studies which used concurrent standard curve quantification (Figure 1d).

**Figure 1:**
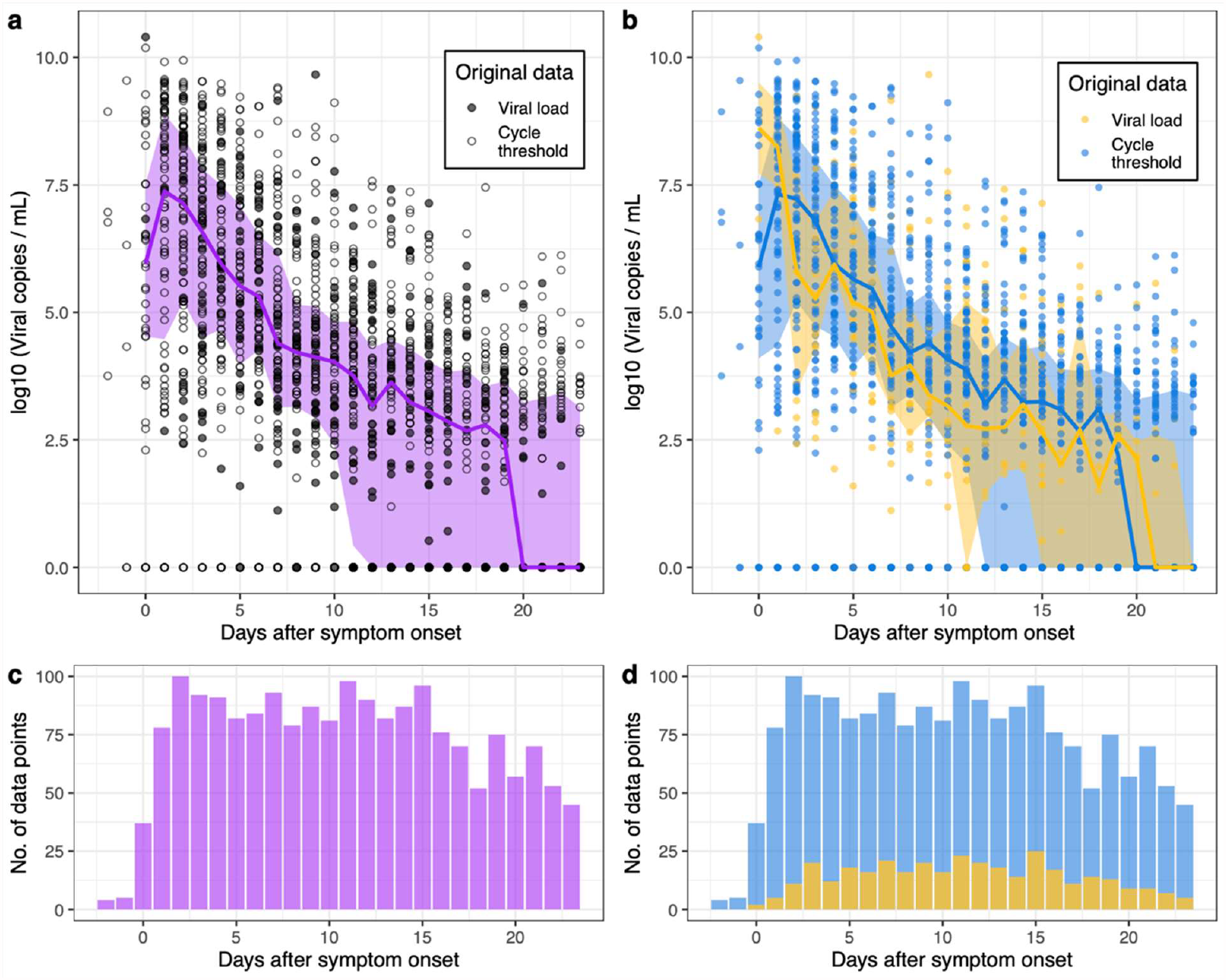
Declining viral loads after symptom onset. a) Data from all 17 studies used in our analysis (circles). For illustrative purposes, viral samples that were negative for virus are set to 1 viral copy per ml. The median viral load is calculated for each day (purple line), as well as the interquartile range (purple shaded region). From Day 20 onwards, over half the samples recorded on each of these days were below the limit of detection. b) Here we show the quantified PCR data (yellow) separately from the data for which viral loads were estimated (blue) using an averaged standard curve (Methods). In the lower panels (c,d), we display the number of data points available on each day for this analysis.

The timing of the first sample obtained relative to onset of symptoms varied with severity of illness. Subjects with moderate or severe disease had first samples collected later in their illness than those with mild disease (Supplementary Figure 3a). Accordingly, first viral load and maximum viral load measurements for subjects with moderate or severe disease in these studies were lower than in those with mild disease (Supplementary Figures 3b and 3c).

### Fitting a regression model to the viral load data

We fitted two types of models to the viral load data from the first 15 days from onset of symptoms (see Methods), using only subjects with at least three samples during this time period (models fitted to 870 samples from 155 patients from 16 studies). The first was a linear regression model, fitted to log-transformed viral loads. This model included patient- and study-specific random effects, which captured variation from the average behaviour observed across 16 datasets. There was variation in the peak and slope of the viral loads across different studies (Figure 2, Supplementary Table 3). The variation in the (log-transformed) peak viral load varied over several orders of magnitude. Some of this variation was explained by study-specific differences: the standard deviation of the between-study variation in the peak viral load was 0.84 i.e. nearly one order of magnitude (Supplementary Table 3). This could be due to a number of factors, such as the method of sample collection, quantification method or characteristics of included patients. Furthermore, viral loads in several studies were estimated using an averaged standard curve, which introduces some uncertainty into the magnitude of the viral load. However, the inclusion of study-specific random effects allows such data to be appraised alongside data from other studies.

**Figure 2:**
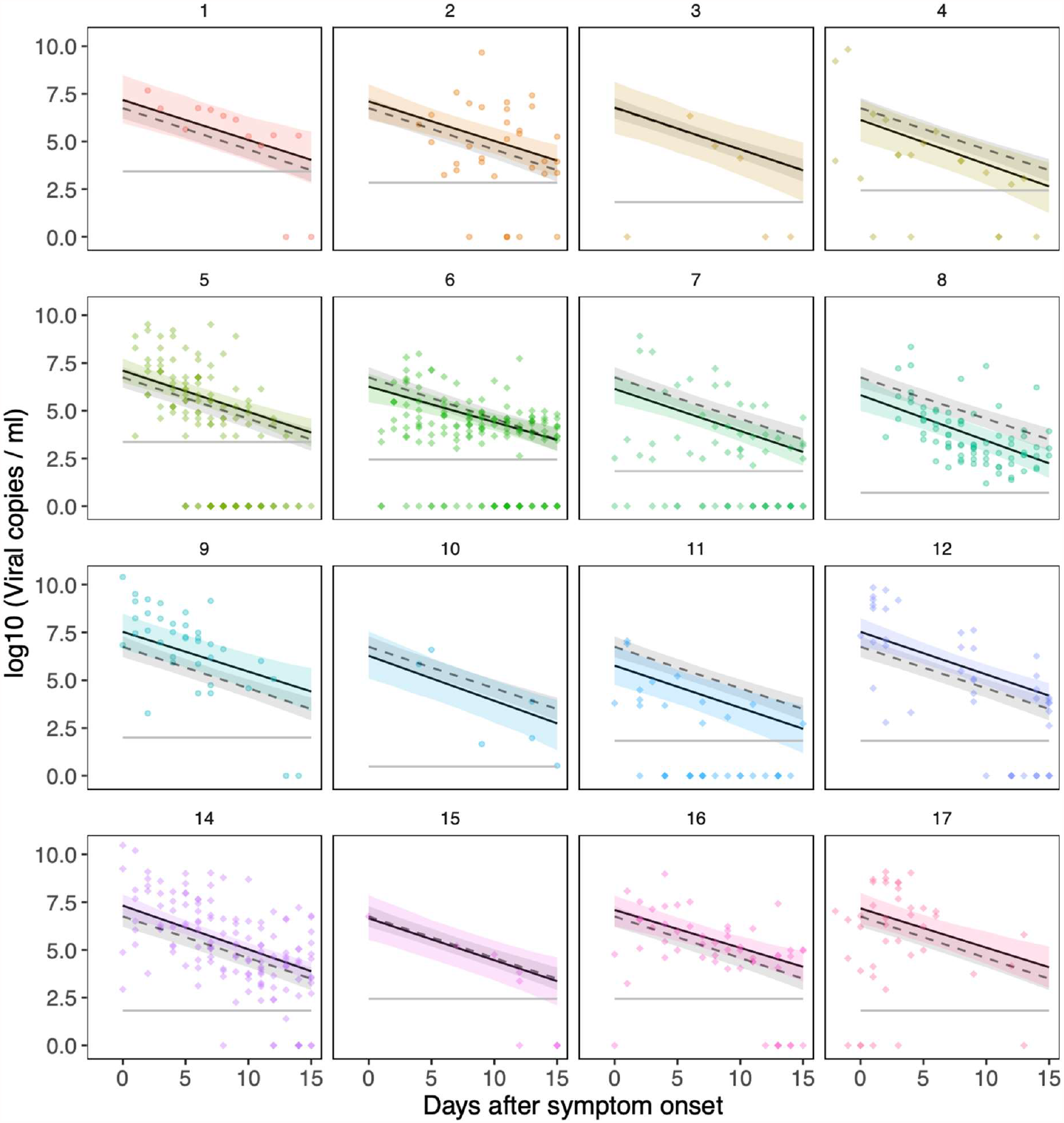
Viral load data and mixed-effects regression model. Data from all 16 studies used in the regression modelling (numbered as in Table 1), showing samples taken within the first 15 days of symptom onset. We fitted a regression model to the data, with study-specific random-effects for the peak viral load and rate of decline (slope). The solid lines show the posterior mean behaviour for each study, with the shaded areas showing the 95% credible intervals. The dashed line, which is the same in each panel, is the average trajectory across the 16 studies. The 95% credible interval for the averaged trajectory is shown by the grey shaded region. Population-level parameters for this model are shown in Supplementary Table 3.

Within this regression framework, we incorporated information on age, disease severity and sex (Methods) to see if the goodness of fit could be improved. We added fixed effects for these three variables, both separately and in combination. We separately examined age as a continuous variable or a dichotomous variable (stipulating whether patients were 60 years of age or over). The goodness of fit to the data did not vary appreciably: here we report results for age as a dichotomous variable. As we had relatively few (29) samples from patients with severe disease in the subset of the data considered here, we pooled patients with moderate disease and severe disease together. The inclusion of the fixed effects for age, sex, or severity did not improve the model fit (Table 2). In other words, the wide variation between individuals observed in the viral load dynamics could not be explained by the inclusion of these variables (Figure 3).

**Table 2:**
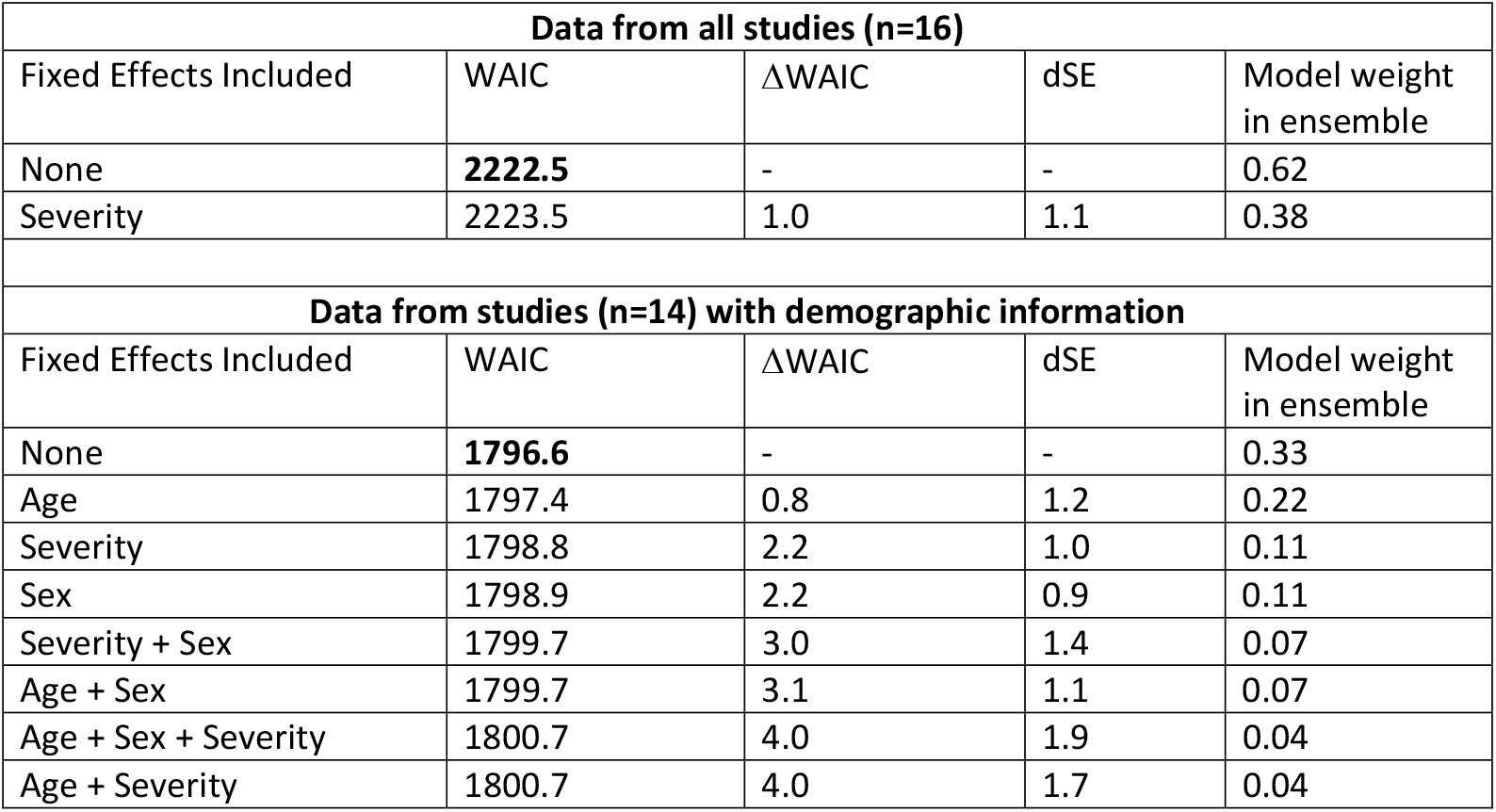
Assessing the goodness of fit of the regression models. Three fixed effects (severity, age, sex) were added to linear regression models of viral load over time, separately or in combinations (see Methods), to determine the extent to which they explained variation present in the data. All studies included in this analysis described the severity of disease for each patient. However, patient-level demographic information was only provided in 15 of the 17 studies. Hence, a separate analysis was carried out for these studies. In each analysis, the best-fitting model is the one with the lowest WAIC value, as indicated in bold. The value of ΔWAIC indicates the difference in goodness of fit between each model and the one that provided the best fit to the data, whilst dSE denotes the standard error of the difference in WAIC between each model and the best-fitting one. In each analysis, we provide the Akaike weight of each model in the ensemble of candidate models that were analysed (Methods).

| Data from all studies (n=16) |  |  |  |  |
| --- | --- | --- | --- | --- |
| Fixed Effects Included | WAIC | ΔWAIC | dSE | Model weight in ensemble |
| None | **2222.5** | - | - | 0.62 |
| Severity | 2223.5 | 1.0 | 1.1 | 0.38 |
| Data from studies (n=14) with demographic information |  |  |  |  |
| Fixed Effects Included | WAIC | ΔWAIC | dSE | Model weight in ensemble |
| None | **1796.6** | - | - | 0.33 |
| Age | 1797.4 | 0.8 | 1.2 | 0.22 |
| Severity | 1798.8 | 2.2 | 1.0 | 0.11 |
| Sex | 1798.9 | 2.2 | 0.9 | 0.11 |
| Severity + Sex | 1799.7 | 3.0 | 1.4 | 0.07 |
| Age + Sex | 1799.7 | 3.1 | 1.1 | 0.07 |
| Age + Sex + Severity | 1800.7 | 4.0 | 1.9 | 0.04 |
| Age + Severity | 1800.7 | 4.0 | 1.7 | 0.04 |

**Figure 3:**
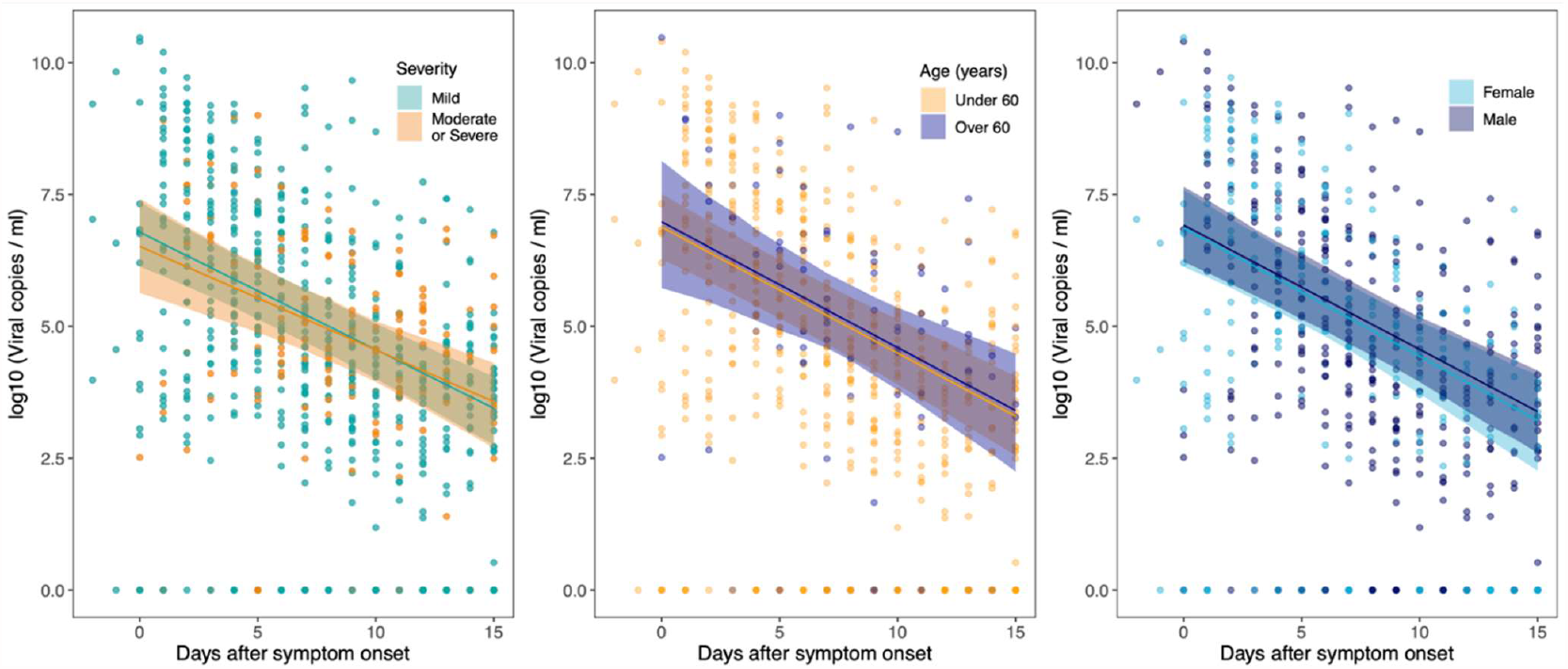
Inclusion of fixed effects in the regression models. All regression models included study- and patient-specific random effects for the peak and slope (i.e. rate of decline) of the viral load. We then added fixed effects, both separately and in combination, to see if the model fit could be improved (Table 2). These fixed effects were: age, sex, and severity of disease. Here we show results for the three models containing one fixed effect (left: severity; middle: age; right: sex). The inclusion of severity, age, or sex did not improve the goodness of fit (Table 2). In the modelling here, age was included as a binary variable (under or over 60 years of age).

We used simulation-based methods (Methods) to estimate the power of our analysis to detect different effect sizes for severity, age and sex on peak viral load (Supplementary Figure 4). For severity and sex, our analysis has 80% power to detect a difference of around 1.1 in the log10 viral load (about a 12-fold difference in viral load), whereas power was lower for detecting the effect of age. Importantly the detectable differences are considerably smaller than the inter-individual variation in viral load at any given time point (Supplementary Table 3), indicating that these are not the major determinants of viral load.

### Fitting a mechanistic model to the viral load data

In addition to modelling viral load decline using regression models, we also developed a mechanistic model which we fitted to the dataset. We elected to keep the model relatively simple due to lack of identifiability between more complex model structures. We represented the multi-faceted immune response to the infection via two components. First, the exponential growth of viral load in the initial phase of infection is brought under control by an *early immune response*. Subsequent to this, the infection is gradually cleared by a *late immune response*. The early immune response is stimulated by a high load of infected cells and starts to block viral replication and the invasion of susceptible cells. The late response requires a maturation phase before it becomes effective and is therefore, more representative of the adaptive immune response. However, we do not attempt to fully distinguish between innate and adaptive responses in this model, as the interplay between them is complex.

To guide the model fitting (see Methods), we made the assumption that both the peak viral load and the activation of the early immune response should roughly be concurrent with the onset of symptoms. The vast majority of subjects in the dataset were only under observation after the onset of symptoms, which means we are unable to infer the rate of exponential growth during the initial phase of the infection. For each subject, we fitted two parameters in the mechanistic model, holding all other parameters fixed (see Methods). One of these free parameters governs the density of infected cells required to activate the early response, while the other determines the rate at which the late response clears infected cells and, therefore, the rate at which the viral load declines. As we did for the regression modelling, we used a nested random effect structure, with study- and patient-specific random offsets for both of these parameters (Methods). In addition, we incorporated data points that fell below the limit of detection in each study by accounting for censoring in the likelihood (Methods).

This approach allowed us to characterise the average time-course of an infection at the population-level i.e. after removing study- and patient-level offsets (Figure 4, Supplementary Table 4), as well as showing the dynamics for the study specific fits to the data. There was no significant relationship between the subject-specific random effects for either of the model parameters and maximum disease severity (Supplementary Figure 5), consistent with the outcome of the preceding regression modelling (Table 2). Similarly, there was no significant relationship between the subject-specific random effects and subject age or sex (not shown). Supplementary Figure 6 shows the 95% credible intervals for the study-specific random effects, showing the between-study variation in both the peak viral load (panel a) and the rate at which the infection is cleared in the URT (panel b). Supplementary Figure 7 shows modelled viral loads for each of the 155 subjects used to fit the mechanistic model.

**Figure 4:**
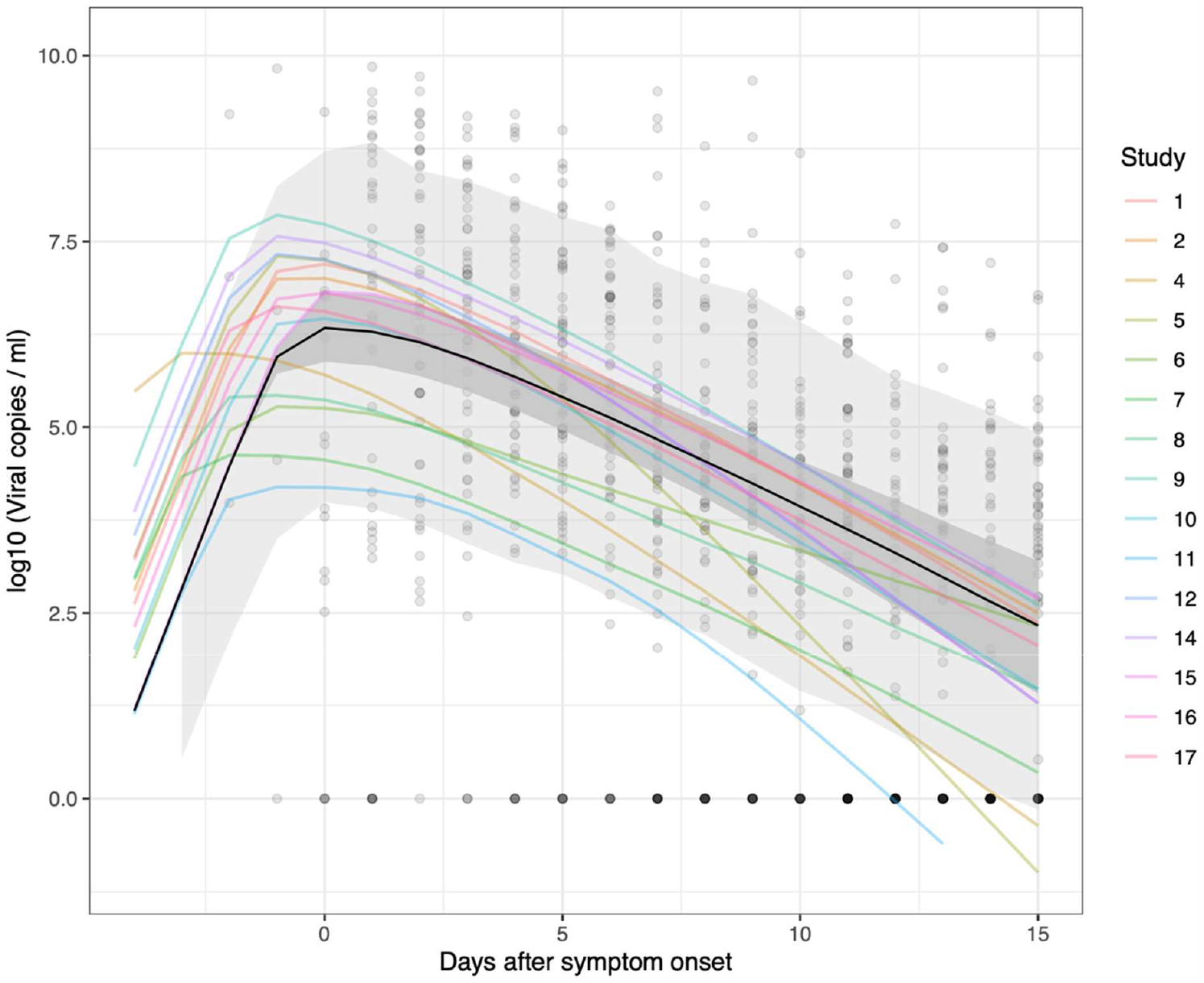
Averaged trajectories obtained from the fitted mechanistic model. In this plot we show the average trajectory predicted for each study (coloured lines), generated using the median value used for the initial viral load at t=0 in each case. We also show the average trajectory across all the studies, indicated by the black line. The dark grey shaded area indicates the 95% credible interval for this average trajectory. The light grey area accounts for the variation observed around the average trajectory (generated using samples from s, as defined in in Equation 8, and calculating the 95% prediction interval for the population-level dynamics). The fit to data from Study 3 is not shown, as this study only contained one patient, which means one cannot distinguish between study- and patient-specific random effects. The opaque black circles are the data points from the 16 studies used to fit the model. For illustrative purposes, viral samples that were negative for virus are set to 1 viral copy per ml (i.e. 0 on the log-scale). The results from the mechanistic model presented here were obtained using 1500 samples from the posterior distribution, with the median trajectories plotted.

### Relating viral load dynamics to SARS-CoV-2-specific immune responses

To demonstrate the potential to link viral load dynamics to SARS-CoV-2-specific adaptive immune responses in individual subjects, we made use of detailed data on neutralising antibody and T-cell responses from the study by Tan et al. (7). We compared this immunological data with the patient-specific late immune response parameter fitted in our mechanistic model in these 12 patients, to attempt to gain insight into within-host mechanisms that might control viral load (Figure 5). Specifically, we assessed the total interferon-γ (IFN-γ) T-cell response to SARS-CoV-2 peptides measured in an ELISPOT assay, and results from a surrogate virus antibody neutralization assay. Simple logistic curves were fitted to the data from each subject’s measured immune responses, and the area under each curve (AUC) was used as a measure of the magnitude of each subject’s immune responses (Figure 5b and 5c). Overall, no significant correlation was observed between the modelled late immune response and the total interferon-γ (IFN-γ) T-cell response (r=0.39, p value = 0.208, see Figure 5d) or the neutralizing antibody response (r=0.07, p value = 0.831, see Figure 5e). However, we noted two subjects (subjects 1 and 12) displayed quite distinct immune responses to the others, characterized by absent neutralizing antibody, and initially high but subsequently declining total interferon-γ (IFN-γ) T-cell responses, possibly indicating a qualitatively different response to SARS-CoV-2. When these two subjects were removed from the analysis, a much stronger correlation was observed between the modelled late immune response and surrogate virus antibody neutralization (r=0.79, p value = 0.006).

**Figure 5:**
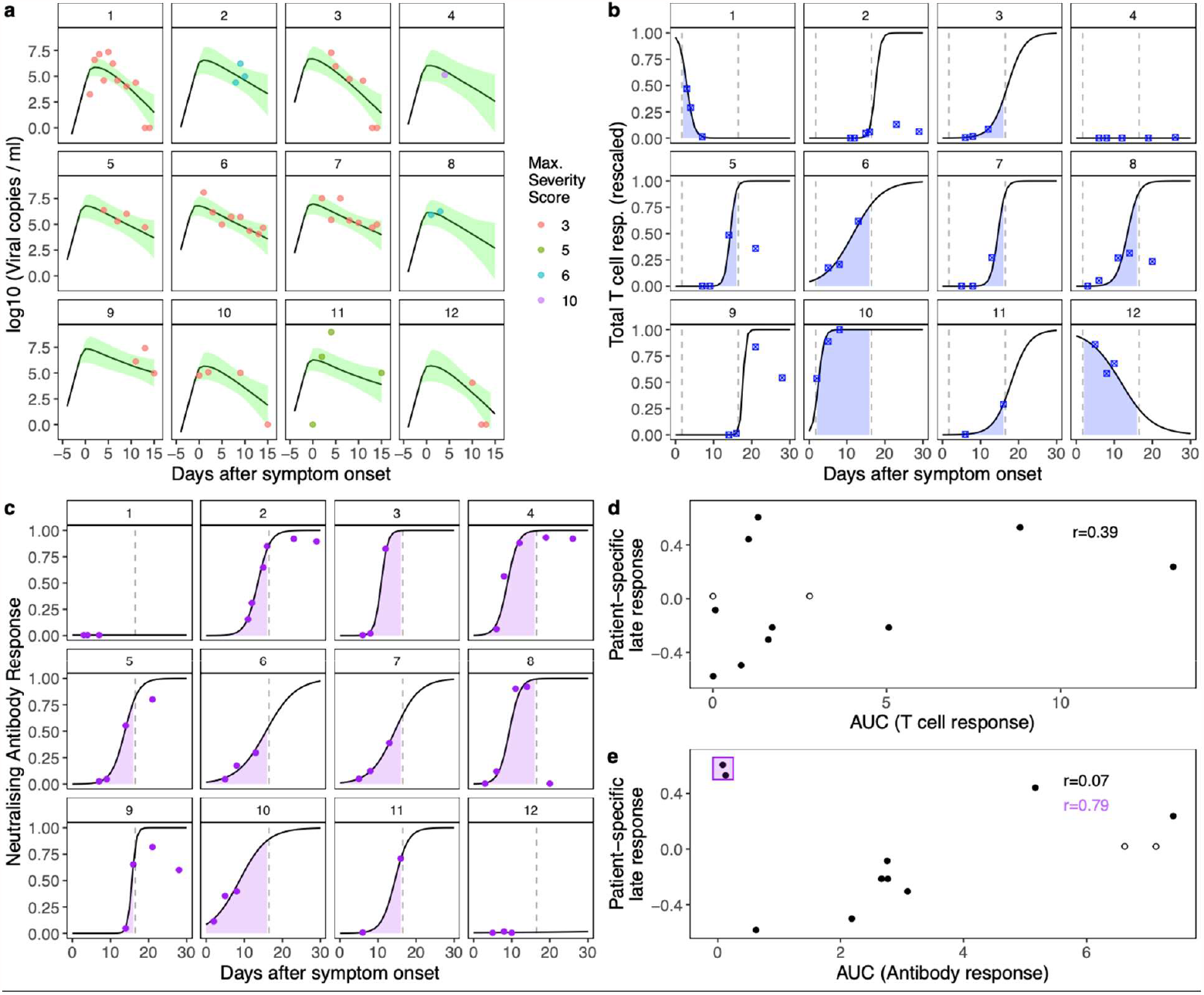
Paired viral load and immune response dynamics. These panels show data from 12 patients, reported by Tan et al. (7). Panel (a): viral load measurements (points) and modelled viral load trajectories from the mechanistic model (black lines show the posterior means, shaded areas are the 95% credible intervals). The coloured symbols indicate the severity score recorded for each subject (on the WHO severity scale). Panel (b): measured total T cell response (blue symbols), rescaled by the largest observed measurement. A logistic curve was fitted through the points for each patient, to facilitate the area under the curve (AUC, blue shaded area) calculation. To calculate both AUCs (antibody & T cell) we used only the first 15 days after symptom onset, as this was the time period used to fit our models. Panel (c): neutralising antibody response (purple dots). A logistic curve was fitted through the points for each patient, to facilitate the area under the curve (AUC, purple shaded area) calculation. Panel (d): relationship between the calculated AUCs of the T cell responses and modelled patient-specific immune responses (p value = 0.208). Panel (e): relationship between the calculated AUCs of the antibody responses and modelled patient-specific immune responses. Two patients failed to mount an antibody response which neutralised virus. The correlation between the patient-specific response and the AUC is much stronger when these patients are not included (p value = 0.006, compared to p value = 0.831 when all 12 subjects are considered). We note that for subjects 4 and 8 (open circles in panels d and e), fewer than 3 viral load measurements were available, meaning their fitted parameters may shrink to the study mean.

## Discussion

Understanding the causes and consequences of variation in pathogen load is fundamental to infectious disease research (35). Increasing pathogen load can drive both pathogenesis and transmission of infection (36). High viral load in COVID-19 has been associated with severity of illness in some studies (3,4), but not others (see e.g. Ref. (37)), and has been associated with risk of transmission (5). Pathogen load is dynamic, it varies over time, and is often considered to be the stimulus for the host response, as well as a target of the host response. Therefore, any attempt to establish the determinants of pathogen load and relationships between pathogen load, severity, and transmissibility, should account for these dynamics.

We collated longitudinal URT viral load data from 2172 samples taken from 605 subjects with SARS-CoV-2 infection in 17 studies, to investigate the association of viral load dynamics with age, sex, and severity of illness. We used the WHO clinical progression scale to standardise varied descriptions of disease severity reported in different studies. We used two distinct modelling approaches to characterise viral load dynamics, accounting for systematic differences in viral load estimation between studies. We found no evidence to support the hypotheses that URT viral load dynamics are substantially influenced by age or sex, or that URT viral load dynamics influence severity of illness. We also found no association between severity and the latent variables describing early and late immune responses to URT virus in our mechanistic model. Nevertheless, we identified considerable inter-individual variation in URT viral load dynamics. Understanding the biological basis for this variation could help to identify new approaches to reduce transmission of SARS-CoV-2.

The lack of association between URT viral load dynamics and severity of illness is particularly interesting. On one hand, this is not necessarily surprising since severe COVID-19 is predominantly a consequence of LRT and systemic disease processes, and some studies have indicated that viral load in LRT samples is more strongly associated with severity of illness (9). On the other hand, this would imply that distinct processes govern URT and LRT viral load dynamics, or that the extension of infection from URT to LRT and other systemic locations is controlled by different mechanisms to those controlling local viral load. Although it is difficult to obtain serial LRT viral load measurements, evaluating distinct mechanisms controlling local and spatial viral dynamics could be important to understand the pathogenesis of COVID-19 and other respiratory infections.

Our study provides one of the most comprehensive assessments of URT viral load dynamics, but despite collating data from a large number of studies we had a relatively low proportion of patients with very severe illness and very few from those with fatal infection, potentially reducing our ability to distinguish different viral dynamics in these groups. We also had very little data on URT viral loads before the onset of symptoms, which limits our ability to model variation in the rate of increase in viral load early in infection. We lacked data on the interval from exposure to symptoms, forcing us to fix this parameter in our mechanistic model. This means we were unable to properly explore the role that the initial viral dose plays in the dynamics. Furthermore, we did not have sufficient data on ethnicity, or other host characteristics beside age and sex, for which it may have been instructive to examine associations with viral load dynamics. We extracted data on antiviral treatment in different studies where possible (Supplementary Table 2), but there were too few subjects receiving each treatment to allow meaningful analysis.

We used two modelling approaches to analyse the data. Mixed-effects regression modelling sought to determine if any of the wide variation observed in patients’ URT viral loads could be explained by age, sex, or disease severity. The inclusion of these variables, separately and in combination, did not improve the model fit. However, our power analysis suggests that we cannot confidently exclude smaller differences in viral load (e.g. an increase or decrease in peak viral load of less than ten-fold) due to these variables. We believe our power analysis provides useful insight into the capacity of such models to detect differences in viral load dynamics in different subpopulations.

In order to utilise as much data as possible, we have included semi-quantitative data (presented as Ct values) alongside fully quantitative viral load data. We have converted the former using an averaged standard curve. Modelling the data with study-specific random effects provides a way to analyse all the data collected, without requiring study-specific standard curve data. Adding data from more studies in future, will allow us to examine whether the conclusions of our analyses are influenced by this approach.

It is interesting to compare our mechanistic model to others that have been fitted to viral load data. Goyal et al. (26) developed a more complex model, which was able to capture a changing rate of decline in viral load observed in the patient data available to them. However, one should note that in our model we are fitting to a narrower timeframe (no more than 15 days after symptom onset, due to lack of viable infective virus after this time), so it may be that the changing rate at which viral load declines is not so noticeable during this phase of the infection. Another difference between the two approaches is that we use the time of symptom onset to centre the dynamics (i.e. set time equal to zero), whereas Goyal et al. used the time of the first swab. When considering data from lots of different subjects, using symptom onset for temporal alignment helps adjust for the wide variation in the time of sample collection, particularly as we observed a relationship between disease severity and the time of the first sample collection in the studies considered here (Supplementary Figure 3). Among the within-host models already published, one finds wide variation in model complexity. Some models, such as the one presented here, have erred on the side of parsimony, whilst others have sought to capture very intricate within-host processes (see e.g. Ref. (31)). An interesting avenue for further work would be to use goodness-of-fit criteria, such as the one used here for the regression modelling, to explore the extent to which more complicated models explain more of the variation present in the data.

We have demonstrated the potential to relate modelled viral load dynamics, and the immunological determinants of the model, to measured immunological data for individual subjects. It is not surprising that circulating SARS-CoV-2-specific T-cell responses are poor correlates of the late immune response controlling viral load, because these cells would need to migrate from the circulation to other locations like the respiratory tract to control virus. It is reassuring that when we considered individuals who did mount a neutralizing antibody response, we saw that antibody neutralization did correlate well with the late immune response parameter of our model, consistent with the evolving evidence that neutralizing antibody does indeed play an important role. However, it is now well established that some individuals do not mount a detectable serum antibody response to SARS-CoV-2, nevertheless they have protective immunity against re-infection (38), and applying this modelling approach to much larger numbers of subjects might help to identify alternative or additional protective mechanisms. Due to the dynamic interplay between viral load and the immune response, more biological insight can be gained from mechanistic modelling, compared to using summary statistics or regression modelling.

Overall, our analyses indicate considerable variation between individuals in the dynamics of URT viral load, but no association between this variation and severity of illness, or with important biological determinants of severity. This has important implications for investigation of the mechanisms driving both pathogenesis and transmissibility of SARS-CoV-2 infection. We present our mechanistic model as a resource for researchers to relate URT viral load dynamics to biological traits, in order to further unravel the mechanistic determinants, and identify possible targets for interventions to reduce URT viral load and prevent transmission.

## Materials and Methods

### Data

To collect data on viral load dynamics, we searched PubMed and medRxiv for studies that recorded longitudinal viral load data from individuals with symptomatic COVID-19 infections. Searches were carried out between May 20^th^ 2020 and February 11^th^ 2021. In particular, we searched for studies which reported the timing of symptom onset for each patient, which we used to temporally align samples from different patients. We identified 53 studies, data from 5 of which could be extracted from the publication or preprint. We contacted the authors of 48 studies by email to request access to patient-level data, and we received data from 14 of these studies. 2 of the 19 studies were dropped from this analysis, as they contained little or no longitudinal data for URT samples. Studies for which data was obtained are summarised in Table 1: all remaining studies identified by our literature search are summarised in Supplementary Table 1. We extracted as much information as possible on the severity of illness experienced by the patients and extracted demographic information on the patients where available. Two authors independently studied the severity information and matched the descriptions to the WHO clinical progression scale (34). Some articles contained detailed information on the course of disease for each patient, but this was not available in all the studies. We considered scores of 1-3 to represent mild disease, 4-6 to represent moderate disease and scores above 6 to represent severe disease. This is slightly different to the patient state descriptors of the WHO scale, because the majority of studies did not provide sufficient detail about the method of oxygen delivery to allow us to distinguish between scores of 5 and 6. In some studies, the severity of symptoms was recorded at multiple timepoints, over the course of the infection. Here, we use the term ‘severity’ to describe the maximum severity of disease experienced by each patient.

Although some studies with a long follow-up period demonstrate that some patients can remain PCR-positive for virus for well over a month (39,40), several studies (16,17) have demonstrated that very few people are infectious, based on URT swabs, beyond 10 or 14 days after symptom onset. Data collected after this period is likely to reflect RNA debris that remains in the URT. Therefore, our models of viral load are fitted to data points within the first 15 days of symptoms. This means that we have fewer data points to fit to (Table 1), but we believe that the meaningful dynamics, in terms of how the host controls the infection, are found in this time period.

### Viral load Quantification

In all studies real-time polymerase chain reaction (PCR) assays were used to quantify URT virus either as Ct values, or as viral copies /mL (*V*). As discussed in Ref. (41), calculation of viral copies per ml from Ct values requires the use of a ‘standard curve’, which is calibrated to the experimental set-up in a particular laboratory using reference samples. In general, these curves have the form:

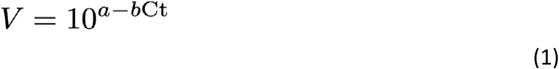

Here *a* and *b* are positive numbers that fully specify the viral load for a given Ct value. This relationship indicates that there is a linear relationship between log-transformed viral loads and the raw Ct values, with higher Ct values representing lower viral loads. The units of *V* vary between studies (e.g. viral copies per ml, per swab, or per 1000 cells), all studies collected here that have quantified viral load have used viral copies per unit of volume. We collected as many different standard curves as possible, from the studies included in this analysis (42,43) and from other papers in the COVID-19 literature (33,44–46), to understand the variation observed. From these, we determined an ‘averaged’ standard curve (Supplementary Figure 1), using the mean observed values for parameters *a* and *b*, which we used to estimate the viral loads for studies for which only Ct values were available. This enabled us to pool data from all 17 studies.

### Models

We sought to explain variation in viral load (either its peak value or its rate of decline over time) among patients, due to (e.g.) age, sex, and severity of disease. We did this using two types of models, a linear mixed effects regression approach and a mechanistic model, which took the form of a system of first-order differential equations. Both models were fitted to viral load data from within 15 days of symptom onset, using only subjects with least three samples taken during this time period (models fitted to 870 samples from 155 patients from 16 studies). Therefore, as indicated in Table 1, no data from study 13 was used to fit the models.

### Linear regression models

Bayesian regression models were fitted using RStan (47), with some of the analysis carried out using the rethinking package (48). Linear models were fitted to log-transformed viral loads, with random effects for each patient and study applied to the parameters determining the peak viral load, which we assumed coincides with the onset of symptoms, and rate of its decline over time. Samples for which no virus was detected were treated as being below limit of detection (LOD), rather than truly virus negative. We show the general form of the regression model here, where *L* is the likelihood of each data point, to illustrate the random-effect structure and the censoring of the data:

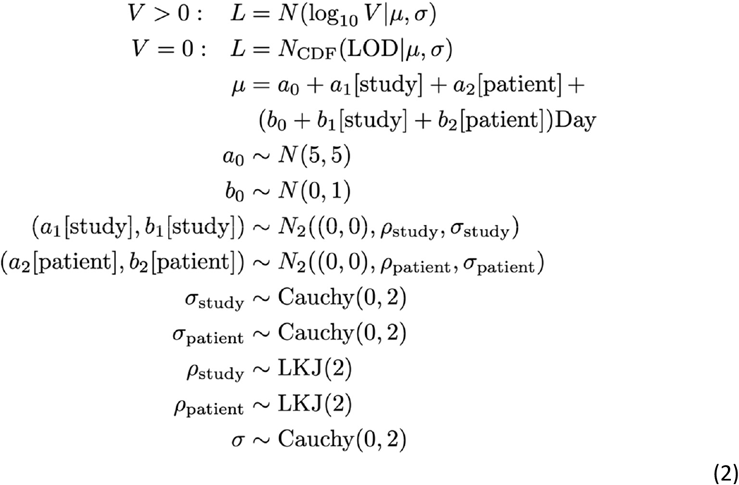

This model is relatively simple: the log-transformed viral loads are captured by a linear model, with the intercept describing the viral load at the time of symptom onset, and the slope capturing the rate at which the viral load subsequently declines. The log-transformed viral load for a given patient is normally distributed around a modelled trajectory, which is described by *μ*, with a standard deviation given by *σ*. For data points where no virus was detected, the likelihood takes a different form. We calculate the likelihood as the probability of the viral load being below the LOD, writing *N*_*CDF*_ as the cumulative distribution function of the normal distribution. We allow for the fact that the LOD is study-specific (indicated in Figure 2). All remaining terms in Equation 2 define the prior distributions for the parameters. Both the slope and the intercept have a parameter that captures the average behaviour across all patients in all studies (*a*_0_ and *b*_0_ respectively). Random effects capture patient- and study-specific deviations from the average behaviour. Both the patient- and study-specific random effects are normally distributed with zero mean: here we write *N*_2_ to indicate the bivariate normal distribution. We allow correlation between the random effects on the intercept and slope at each level, as indicated by the presence of parameters *ρ*_*study*_ and *ρ*_*patient*_. For these parameters, we use the LKJ distribution (49) as a prior.

Within this framework, fixed effects could then be added to the model, to assess whether (e.g.) severity of disease, or the sex of patient affected the typical viral load trajectory observed. To do this, we simply add terms to *μ* in Equation 1. Here we illustrate this for sex, making female the reference category:

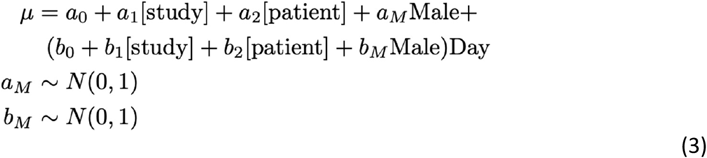

Here variable ‘Male’ is equal to 1 if the patient is male (0 otherwise), and we have also added zero-mean priors for the new parameters. As mentioned above, severity was expressed on the WHO scale, which takes values from 1-10. As 1 is asymptomatic, severity in our dataset is limited from 2 (mild symptoms, not hospitalised) to 10 (dead). As the dataset contains relatively few samples from patients with severe disease (score 7 to 10 on the WHO scale), we chose to make two groups from these categories: Mild (2 or 3) and Moderate or Severe (4-10). When severity is included in the model, *μ* becomes:

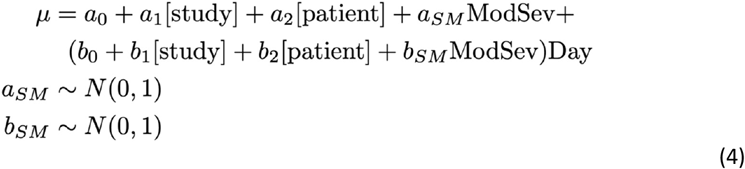

Here variable ‘ModSev’ is equal to 1 for patients with moderate or severe disease (0 otherwise). In most studies, patient age is given in years, as an integer value. Some studies expressed age of patients in decades e.g. 10-20 years of age. For these studies, we used the midpoint of the age range given for each patient. We grouped age into two groups: _ <60 years of age and ≥60 years of age. In the regression modelling, we added a fixed effect for age, making the youngest age group the default. Here, *μ* takes the form:

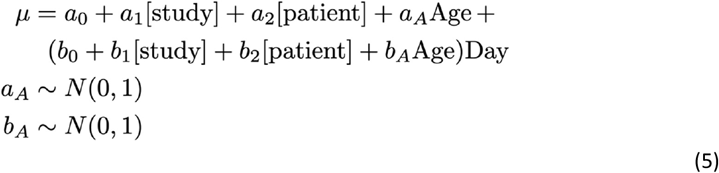

Here variable ‘Age’ is equal to 1 for patients aged 60 or over (equal to 0 otherwise). We also looked at including subject age as a continuous variable, but the goodness of fit did not change appreciably. We added the 3 terms (age, severity, sex) to the regression models both separately and in combination. We assessed the goodness of fit using the Watanabe Akaike Information Criterion (WAIC) (49), with the best-fitting model having the lowest WAIC value. As the WAIC for each candidate model is estimated from a finite sample, its standard error was calculated using the rethinking package (48) to appreciate the uncertainty in its value. These standard errors are useful when appraising the differences in WAIC between candidate models. The relative goodness of fit of a given model can also be appraised by calculating its Akaike weight among the set of all considered models. This can be interpreted as the probability that this model, out of the set of models considered, would provide the best fit to new (i.e. out of sample) data (49).

We use simulation-based estimation of statistical power to assess our capacity to detect a difference in viral load dynamics due to one of the three factors (sex, age, severity of disease) assessed here. We generated synthetic datasets of the same size as the one considered here, with study- and patient-specific random effects of the same magnitude (Supplementary Table 3). For simplicity, we generated datasets with an equal number of samples per subject, and the sampling times were randomly generated from a uniform distribution. When generating the data, we assumed that the peak viral load was influenced by one of the three aforementioned factors. We then ran the regression analysis, to see if the modelled effect could be detected. To reduce computation time, we here used frequentist regression via the lme4 package in R (50), with a p value < 0.05 for the included fixed effect indicating a significant finding. Generating 1000 synthetic datasets, the statistical power can be estimated as the percentage of datasets for which the regression analysis found a significant effect. Supplementary Figure 4 shows how the statistical power varies with the magnitude of the modelled effect, which is here expressed as a fold difference in the peak viral load. The results of the power analysis suggest that we were not well powered to measure relatively small differences in peak viral load (under 10-fold difference). The power to measure sex-specific differences in peak viral load was slightly higher than the power to measure severity- or age-specific differences, as we have a better balance between samples from male and female samples than we do between samples in under 60s and over 60s, or samples from subjects from mild disease versus samples from subjects with moderate or severe disease.

### Mechanistic model

We also developed a mechanistic model to describe the viral dynamics. We started modelling the infection 5 days before the onset of symptoms. At this time, an initial dose of virus *V*_0_ is introduced. These virions start invading susceptible cells at rate *β*. Infected cells then produce more virus at rate *p*, which can then invade subsequent susceptible cells. Meanwhile, free virions are cleared at rate *γ*. In this way, the viral population grows exponentially. In the model, this growth is brought under control by an early phase of the immune response, and then the infection is gradually cleared by a later phase of the immune response. We do not seek to specify the immunological mechanisms contributing to these phases of the immune response. In each infection, the exponential growth slows as the population of infected cells approaches a certain value (*I*_*max*_). This reflects a combination of two within-host mechanisms: the depletion of susceptible cells in the URT and the effect of an early phase immune response which is triggered at a high level of infection. Since these mechanisms may be linked, i.e. the immune response may modify the susceptibility of target cells, we do not attempt to distinguish between them, or to explicitly model the population of susceptible cells.

After the exponential growth is brought under control, the infection is cleared by the late immune response. In this model, this is triggered by a certain density of infected cells but requires a maturation stage before it becomes effective at clearing infected cells. We write the system of equations as:

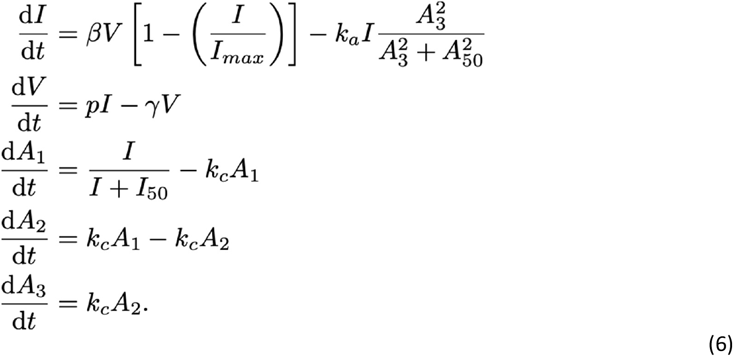

The late immune response, represented here by variables (*A*_1_, *A*_2_, *A*_3_), is stimulated by the presence of infected cells. A Hill function is used to make the adaptive response dimensionless, and to rescale its value (daily input into compartment *A*_1_ scales between 0 and 1). Parameter *I*_50_ (here fixed to a value of 1000 cells) determines the magnitude of the density of infected cells required to stimulate this response. It requires time to mature (governed by rate parameter *k*_*c*_, which we fix at 0.33day^-1^), meaning that only stage *A*_3_ is able to clear infected cells. Once mature, the late response does not wane in this model, as we are only interested in its ability to clear an infection, not how long it persists after the infection has been resolved. In this way the late response recapitulates features of adaptive cell-mediated and humoral immunity, without needing to specify their relative contributions. Parameter *k*_*a*_ represents the maximum rate at which the late response can clear the infection, and *A*_50_ governs the magnitude of the adaptive response required effectively clear infected cells. We note that the magnitude of the late immune response has been set to be of order 1 for convenience, and this determines the magnitude of *A*_50_.

For the vast majority of patients for whom we have data, only samples taken after the onset of symptoms are recorded. This means that we are unable to estimate the duration of the incubation period, the initial dose of virus that causes the infection, or the rate at which the virus reproduces before being acted upon by the immune response. We elect to model a five-day incubation period and fix the rate of exponential growth to be the same for all patients, setting *β* = 0.8 ml day^-1^virion^-1^, *γ* = 13day^-1^, *p* = 80day^-1^, with these values informed by parameter values chosen in published models (25,26,28). This is a simplification: it is unclear whether, in reality, the time from infection to symptom onset varies by age, sex or disease severity. For each patient the infection starts at Day -5 (in the dataset, Day 0 is the day of symptom onset, not the day of infection onset) with initial conditions given by (*I* = 0, *V* = *V*_0_, *A*_1_ = 0, *A*_2_ = 0, *A*_3_ = 0) i.e. the infection starts with an initial viral dose, no infected cells, and no prior exposure to the virus (late immune response completely inactive).

The initial viral dose, 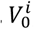, for each patient is chosen with the idea in mind that the viral load should peak at or close to the time of symptom onset. Ideally, 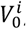, should be fitted simultaneously with the free parameters for each patient, but this has proved challenging to date, especially given that peak viral load should coincide approximately with symptom onset but may also not be observed. Briefly, 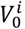, is chosen by considering the linear subsystem of equations that govern the growth of the virus prior to the activation of the immune response to the infection. As this system of equations is linear, it can be solved exactly (we use a matrix exponential). We require that 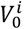 be chosen so that, just prior to the onset of symptoms, an uncontrolled infection would reach the maximum viral load observed for patient *i* in the dataset. This ensures that the peak viral load modelled for each patient is controlled by the early-stage immune response, as it is too soon for the adaptive response to be effective at clearing infected cells.

When fitting to the data, we allowed the values of *k*_*a*_ and *I*_*max*_ to vary and we fixed all the other parameters. As we did for the regression modelling, we included random effects in the model, to account for different behaviour in different patients and across different studies. In the case where we have P patients taken from S studies, we write:

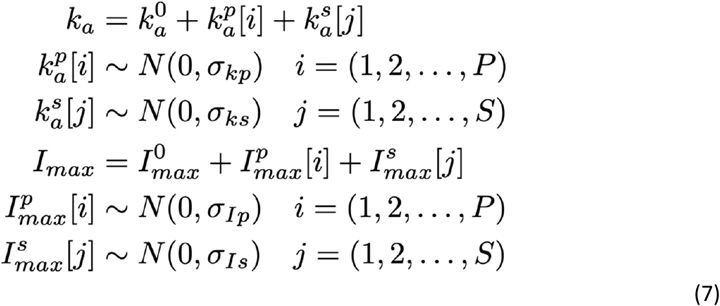

This means that 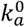 and 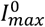 represents the population-level average value of each parameter. The system of equations was solved numerically in R, using the dopri function from the dde package (51). We write the modelled viral load trajectory at time *t* for patient *i* in study *j* as *V*_*ij*_(*t*). Samples from the posterior distribution of the fitted parameter were obtained using Markov Chain Monte Carlo methods, via the R package drjacoby (52). As with the regression modelling, the form of the likelihood for each data point depends on whether the sample is above the study-specific LOD or is recorded as being virus-negative. We write *D*_*ijk*_ to indicate the k^th^ viral load sample, taken *t*_*k*_ after the onset of symptoms, from patient *i* in study *j*. The likelihood for each data point has the form:

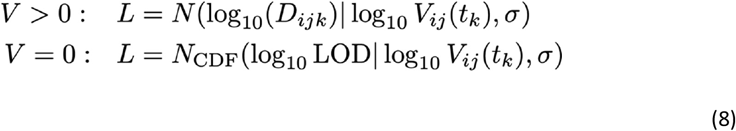

The global likelihood was then obtained by multiplying together the likelihoods for each data point. We provided prior distributions for the parameters to be fitted. These were:

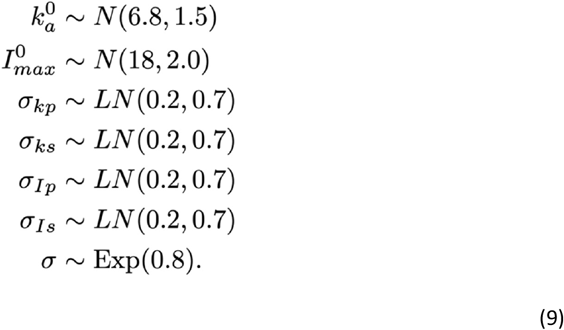

## Supporting information

The Supplementary Materials contain Supplementary Tables 1-4 and Supplementary Figures 1-7

## Data Availability

At present, we refer the reader back to the original studies (summarised in Table 1) to access or request the viral load data.

## Acknowledgments

We would like to express our gratitude to the study authors who shared their data, as well as all those involved in the data collection. In addition, we acknowledge Professor Antonio Bertoletti and colleagues for making subject-level immunology data available. JDC would like to thank Peter Winskill, Robert Verity, and Clare McCormack for useful discussions.

## Supplementary Materials

The Supplementary Materials contain Supplementary Tables 1-4 and Supplementary Figures 1-7.

## Availability of Data

Anonymised viral load data is available alongside this article.

## Availability of Code

The R scripts used to carry out the analyses presented here and generate the figures is available alongside this article. We also demonstrate how the models could be fitted to additional viral load data, if supplied by the reader.

