## Supplementary material for "Modelling upper respiratory viral load dynamics of SARS-CoV-2": The Supplementary Materials contain Supplementary Tables 1-4 and Supplementary Figures 1-7

**Corresponding author*

### SUPPLEMENTARY MATERIALS

This document contains:

1. Supplementary Tables 1-4
2. Supplementary Figures 1-7

*All references numbered as per the main text*

| Title | Journal | D.O.I. | Date of request |
|---|---|---|---|
| Temporal dynamics in viral shedding and transmissibility of COVID-19 | Nature Medicine | https://doi.org/10.1038/s41591-020-0869-5 | May 26th 2020 |
| Temporal profiles of viral load in posterior oropharyngeal saliva samples and serum antibody responses during infection by SARS-CoV-2: an observational cohort study | Lancet ID | https://doi.org/10.1016/S1473-3099(20)30196-1 | Oct 22nd 2020 |
| Prolonged presence of SARS-CoV-2 viral RNA in faecal samples | Lancet G Hep | https://doi.org/10.1016/S2468-1253(20)30083-2 | June 1st 2020 |
| Clinical Course and Outcomes of Patients with Severe Acute Respiratory Syndrome Coronavirus 2 Infection: a Preliminary Report of the First 28 Patients from the Korean Cohort Study on COVID-19 | J Korean Med Sci | https://doi.org/10.3346/jkms.2020.35.e142 | June 1st 2020 |
| Viral Kinetics and Antibody Responses in Patients with COVID-19 | medRxiv | https://doi.org/10.1101/2020.03.24.20042382 | May 26th 2020 |
| Different longitudinal patterns of nucleic acid and serology testing results based on disease severity of COVID-19 patients | Emerging Microbes & Infections | https://doi.org/10.1080/22221751.2020.1756699 | June 1st 2020 |
| The presence of SARS-CoV-2 RNA in the feces of COVID-19 patients | Journal of Medical Virology | https://doi.org/10.1002/jmv.25825 | Oct 21st 2020 |
| Chronological Changes of Viral Shedding in Adult Inpatients with COVID- 19 in Wuhan, China | Clinical Infectious Diseases | https://doi.org/10.1093/cid/ciaa631 | June 1st 2020 |
| Correlation between viral RNA shedding and serum antibodies in COVID-19 patients | Clinical Microbiology and Infection | https://doi.org/10.1016/j.cmi.2020.05.022 | June 1st 2020 |
| Viral load dynamics and disease severity in patients infected with SARS-CoV-2 in Zhejiang province, China, January-March 2020: retrospective cohort study | BMJ | https://doi.org/10.1136/bmj.m1443 | June 1st 2020 |
| Clinical features and dynamics of viral load in imported and non-imported patients with COVID-19 | International Journal of Infectious Diseases | https://doi.org/10.1016/j.ijid.2020.03.022 | June 1st 2020 |
| Antibody responses to SARS-CoV-2 in patients of novel coronavirus disease 2019 | Clinical Infectious Diseases | https://doi.org/10.1093/cid/ciaa344 | August 20th 2020 |
| Phenotype of SARS-CoV-2-specific T-cells in COVID-19 patients with acute respiratory distress syndrome | medRxiv | https://doi.org/10.1101/2020.04.11.20062349 | June 16th 2020 |
| Shedding of infectious virus in hospitalized patients with coronavirus disease-2019 (COVID-19): duration and key determinants | medRxiv | https://doi.org/10.1101/2020.06.08.20125310 | June 16th 2020 |

| | | | |
|---|---|---|---|
| SARS-CoV-2 viral load predicts COVID-19 mortality | Lancet Resp Med | https://doi.org/10.1016/S2213-2600(20)30354-4 | August 14th 2020 |
| Duration of viral shedding in asymptomatic or mild cases of novel coronavirus disease 2019 (COVID-19) from a cruise ship: A single-hospital experience in Tokyo, Japan | International Journal of Infectious Diseases | https://doi.org/10.1016/j.ijid.2020.06.020 | August 20th 2020 |
| Relative COVID-19 viral persistence and antibody kinetics | MedRxiv | https://doi.org/10.1101/2020.07.01.20143917 | August 19th 2020 |
| Kinetics of viral load and antibody response in relation to COVID-19 severity | The Journal of Clinical Investigation | https://doi.org/10.1172/JCI138759 | August 20th |
| SARS-CoV-2 RT-PCR profile in 298 Indian COVID-19 patients: a retrospective observational study | MedRxiv | https://doi.org/10.1101/2020.06.19.20135905 | August 19th 2020 |
| Viral load dynamics in transmissible symptomatic patients with COVID-19 | MedRxiv | https://doi.org/10.1101/2020.06.02.20120014 | August 19th 2020 |
| Clinical Course and Molecular Viral Shedding Among Asymptomatic and Symptomatic Patients With SARS-CoV-2 Infection in a Community Treatment Center in the Republic of Korea | JAMA | https://doi.org/10.1001/jamainternmed.2020.3862 | August 19th 2020 |
| Duration of infectiousness and correlation with RT-PCR cycle threshold values in cases of COVID-19, England, January to May 2020 | Eurosurvellance | https://doi.org/10.2807/1560-7917.ES.2020.25.32.2001483 | August 19th 2020 |
| Clinical Characteristics and Viral RNA Detection in Children With Coronavirus Disease 2019 in the Republic of Korea | JAMA Pediatr | https://doi.org/10.1001/jamapediatrics.2020.3988 | Sept 4th 2020 |
| Temporal profile and determinants of viral shedding and of viral clearance confirmation on nasopharyngeal swabs from SARS-CoV-2-positive subjects: a population-based prospective cohort study in Reggio Emilia, Italy | BMJ Open | http://dx.doi.org/10.1136/bmjopen-2020-040380 | Sept 9th 2020 |
| Viral Dynamics and Immune Correlates of Coronavirus Disease 2019 (COVID-19) Severity | Clinical Infectious Diseases | https://doi.org/10.1093/cid/ciaa1280 | Oct 16th 2020 |
| Clinical Performance of SARS-CoV-2 Molecular Tests | J Clin Microbiology | https://doi.org/10.1128/JCM.00995-20 | Oct 21st 2020 |
| Viral dynamics in mild and severe cases of COVID-19 | Lancet ID | https://doi.org/10.1016/S1473-3099(20)30232-2 | Oct 22nd 2020 |
| Clinical and epidemiological features of COVID-19 family clusters in Beijing, China | Journal of Infection | https://doi.org/10.1016/j.jinf.2020.04.018 | Oct 22nd 2020 |
| Clinical and epidemiological features of 36 children with coronavirus disease 2019 (COVID-19) in Zhejiang, China: an observational cohort study | Lancet ID | https://doi.org/10.1016/S1473-3099(20)30198-5 | Oct 22nd 2020 |

| | | | |
|---|---|---|---|
| Viral loads in throat and anal swabs in children infected with SARS-CoV-2 | Emerging Microbes & Infections | https://doi.org/10.1080/22221751.2020.1771219 | Oct 22nd 2020 |
| Factors of Severity in Patients with COVID-19: Cytokine/Chemokine Concentrations, Viral Load, and Antibody Responses | AJTMH | https://doi.org/10.4269/ajtmh.20-1110 | Nov 26th 2020 |
| The kinetics of viral load and antibodies to SARS-CoV-2 | Clinical Microbiology and Infection | https://doi.org/10.1016/j.cmi.2020.08.043 | Nov 26th 2020 |
| The Correlation Between Clinical Features and Viral RNA Shedding in Outpatients With COVID-19 | Open Forum Infectious Diseases | https://doi.org/10.1093/ofid/ofaa331 | Nov 26th 2020 |
| Molecular and serological characterization of SARS-CoV-2 infection among COVID-19 patients | Virology | https://doi.org/10.1016/j.virol.2020.09.008 | Nov 26th 2020 |

**Supplementary Table 1**: Summary of studies which were identified during the literature search but did not provide data when the corresponding authors were contacted.

| Study | Reference | No. of Patients | Country | Healthcare setting | Recruitment period | Antiviral treatment, n (%) | | Immunomodulatory treatment, n(%) | |
|---|---|---|---|---|---|---|---|---|---|
| 1 | Kim JY *et al.*, J Korean Med Sci, 2020 [42] | 2 | Korea | Hospital | January 2020 | Lopinavir/Ritonavir | 2 (100) | 0 | 0 |
| 2 | Lui G *et al.*, J Infect, 2020 [53] | 11 | Hong Kong | Multicentre, Hospital | February 2020 | Lopinavir/Ritonavir<br>Ribavirin<br>Beta-interferon | 11 (100)<br>8 (73)<br>5 (56) | Hydrocortisone | 1 (9) |
| 3 | Scott S *et al.*, Clin Infect Dis, 2020 [54] | 1 | USA | Community | January 2020 | None used | | None used | |
| 4 | Kim SE, Int J Infect Dis, 2020 [55] | 3 | Korea | Tertiary Hospital | February – April 2020 | Lopinavir/Ritonavir | 1 (33) | None used | |
| 5 | Gautret P *et al.*, Int J Antimicrob Agents, 2020 [56] | 19 | France | Tertiary Hospital | March 2020 | None used | | Hydroxychloroquine | 18 (95) |
| 6 | Young B *et al.*, JAMA, 2020 [57] | 18 | Singapore | Multicentre, Tertiary Hospital | January – February 2020 | Lopinavir/Ritonavir | 5 (28) | None used | |
| 7 | The COVID–19 Investigation team, Nat Med, 2020 [58] | 12 | USA | Multicentre, Community and Hospital | January 2020 | Remdesivir | 3 (25) | Corticosteroids | 2 (17) |
| 8 | Wölfel R *et al.*, Nature, 2020 [59] | 9 | Germany | Hospital | January 2020 | Not reported | | Not reported | |
| 9 | Vetter P *et al.*, mSphere, 2020 [60] | 5 | Switzerland | Hospital | February 2020 | Lopinavir/Ritonavir | 1 (20) | None used | |
| 10 | Lavezzo E *et al.*, Nature, 2020 [43] | 37 | Italy | Community and Hospital | February – March 2020 | Not reported | | Not reported | |
| 11 | Xu Y *et al.,* Nat Med, 2020 [61] | 6 | China | Paediatric cohort, Tertiary Hospital | January – February 2020 | Alpha-interferon | 6 (100) | IVIG | 1 (17) |
| 12 | Shrestha N *et al.*, Clin Infect Dis, 2020 [62] | 230 | USA | Healthcare worker cohort, non–hospitalized | March – April 2020 | None used | | None used | |
| 13 | Fajnzylber J *et al.*, Nat Commun, 2020 [63] | 64 | USA | Multicentre, Tertiary Hospital | NK | Remdesivir | 16 (25) | None used | |
| 14 | Yilmaz A et *al.*, J Infec Dis, 2020 [64] | 54 | Sweden | Tertiary Hospital | February – April 2020 | Not reported | | Not reported | |
| 15 | Alsharrah *et al.*, J Med Virol, 2020 [65] | 29 | Kuwait | Paediatric cohort, Tertiary Hospital | February – April 2020 | None used | | None used | |
| 16 | Tan A *et al.*, Cell Reports, 2020 [7] | 12 | Singapore | Tertiary Hospital | NK | Not reported | | Not reported | |
| 17 | Salvatore PP *et al.*, Clin Infec Dis, 2020 [66] | 93 | USA | Community | March – May 2020 | None used | | None used | |

**Supplementary Table 2**. Summary of antiviral and immunomodulatory treatment in the studies included in analysis.

| Parameter | Interpretation | Posterior Mean | 95% CrI |
|---|---|---|---|
| $a_0$ | Average value of the peak $\log_{10}$(VL) | 6.74 | (6.17,7.30) |
| $b_0$ | Average value of the rate of decline (per day) of the $\log_{10}$(VL) | -0.22 | (-0.26,-0.17) |
| $\sigma_{patient}[1]$ | Standard deviation of the between-patient variation in the peak $\log_{10}$(VL) | 1.54 | (1.26,1.85) |
| $\sigma_{patient}[2]$ | Standard deviation of the between-patient variation rate of decline (per day) of the $\log_{10}$(VL) | 0.15 | (0.12,0.19) |
| $\rho_{patient}$ | Correlation between $\sigma_{patient}[1]$ and $\sigma_{patient}[2]$ | -0.86 | (-0.92,-0.76) |
| $\sigma_{study}[1]$ | Standard deviation of the between-study variation in the peak $\log_{10}$(VL) | 0.81 | (0.42,1.35) |
| $\sigma_{study}[2]$ | Standard deviation of the between-study variation rate of decline (per day) of the $\log_{10}$(VL) | 0.04 | (0.01,0.09) |
| $\rho_{study}$ | Correlation between $\sigma_{study}[1]$ and $\sigma_{study}[2]$ | -0.18 | (-0.80,0.45) |
| $\sigma$ | Standard deviation of the observed variation in $\log_{10}$(VL) around the linear model | 1.08 | (1.01,1.16) |

**Supplementary Table 3**: Summary of the population-level (i.e. not study- or patient-specific) parameter values (and 95% credible intervals) obtained for the multi-level regression modelling (as displayed in Figure 2). Patient- and study-specific random effects were used for both the peak (log-transformed) viral load, and its rate of decline per day.

| Parameter | Interpretation | Posterior Mean | 95% CrI |
|---|---|---|---|
| $k_a^0$ | Population-level late immune response | 6.32 | (5.84-6.74) |
| $\sigma_{kp}$ | Standard deviation of patient-level offset in the late immune response | 13.62 | (12.40-15.02) |
| $\sigma_{ks}$ | Standard deviation of study-level offset in the late immune response | 0.74 | (0.49-1.04) |
| $I_{max}^0$ | Population-level late immune response | 0.66 | (0.31-1.12) |
| $\sigma_{Ip}$ | Standard deviation of patient-level offset in the early immune response | 2.81 | (2.37-3.26) |
| $\sigma_{Is}$ | Standard deviation of study-level offset in the early immune response | 2.85 | (1.92-4.10) |
| $\sigma$ | Standard deviation of variation of viral load around modelled trajectory | 3.21 | (3.00-3.43) |

**Supplementary Table 4:** Summary of the population-level (i.e. not study- or patient-specific) parameter values (and 95% credible intervals) obtained for the mechanistic viral load model (Equations 6-8). Samples from $k_a^0$ and $I_{max}^0$ were used to generate the black line and dark grey shaded area in Figure 4.

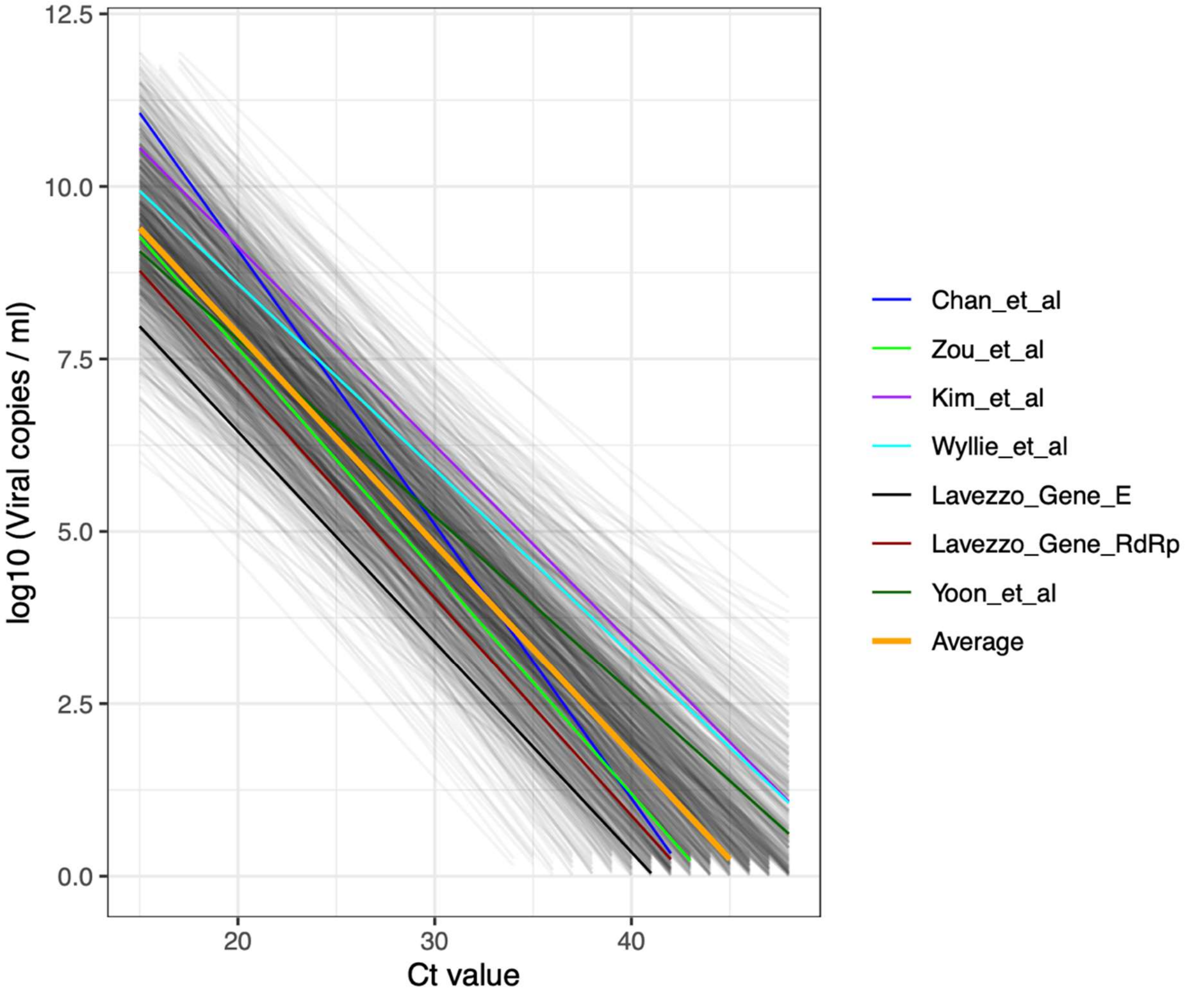


**Supplementary Figure 1**: Standard curves relating cycle-threshold (Ct) values to viral load. Seven standard curves, identified from published studies (see Methods) are plotted. We used these standard curves to produce a model of an averaged standard curve (thicker orange line), and capture the variation observed across different studies. We only used standard curves which quantified viral load in units of viral copies per ml. Standard curves generated by drawing random samples from this model are shown by opaque, grey lines, indicating the potential variation in the standard curve.

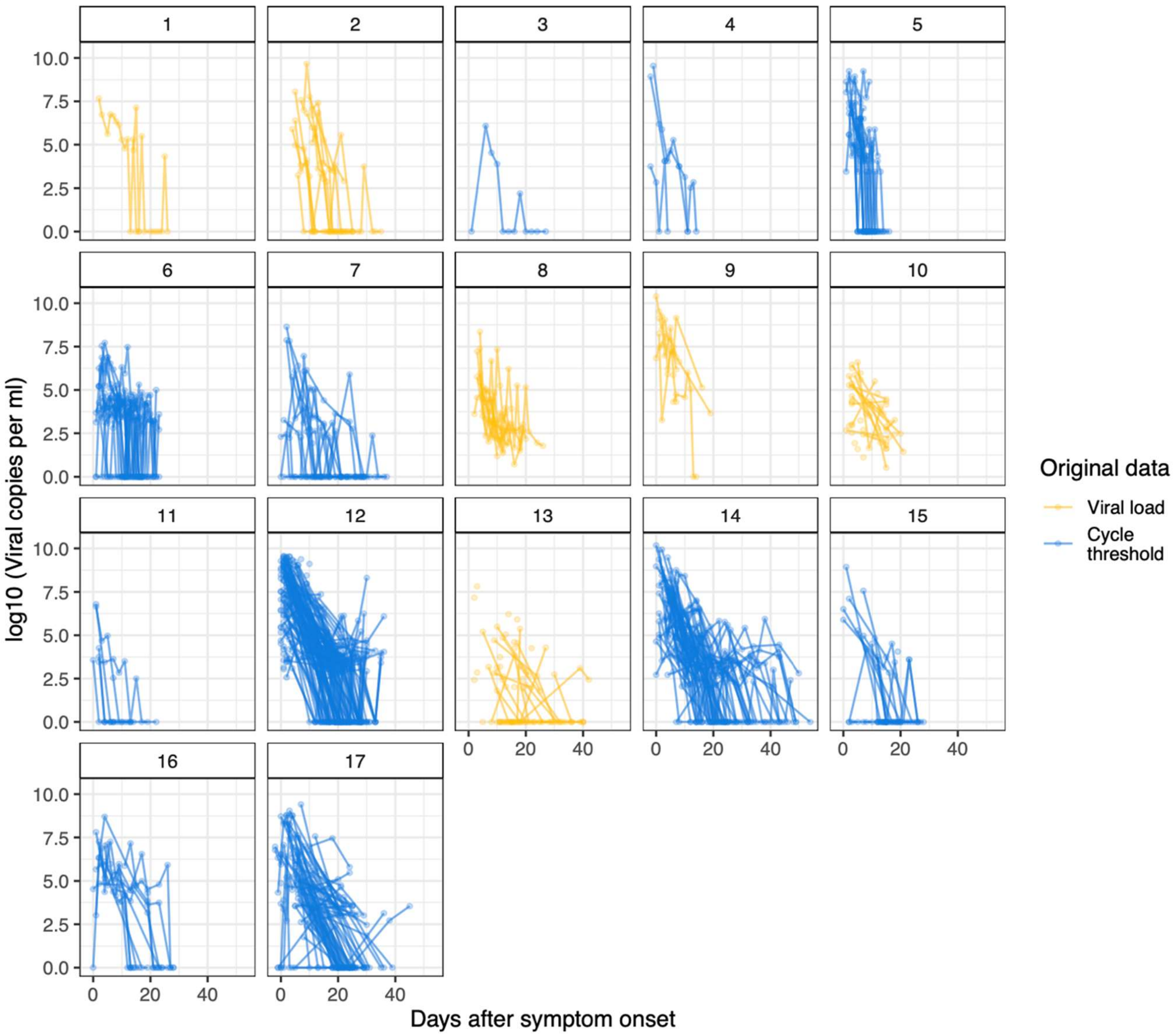


**Supplementary Figure 2**: Summary of all the data collected (see Table 1 in the main text). For the studies shown in blue, viral loads have been estimated using an averaged standard curve (see Methods for details). For illustrative purposes, samples that were negative for virus are set to 1 viral copy per ml. In each panel, the lines connect samples collected from the same patient.

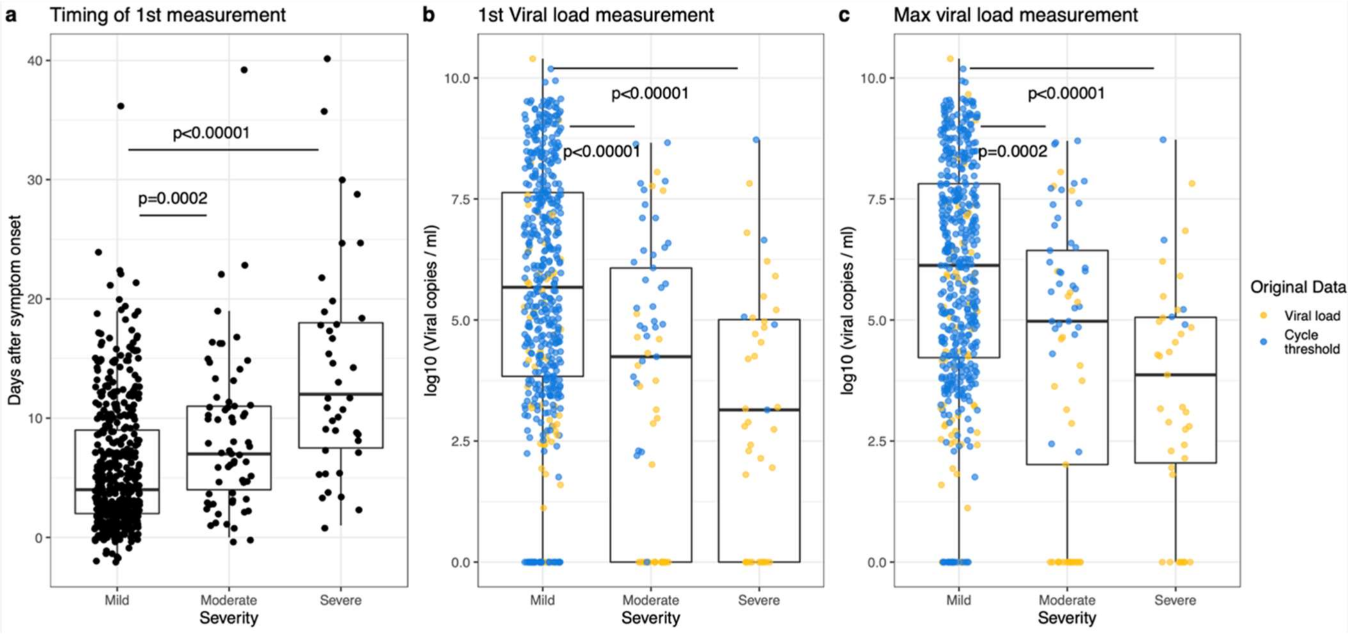


**Supplementary Figure 3**: Comparison of timing of first sample and viral load by severity. a) Timing of first viral load measurement for each patient, relative to symptom onset. Here severity indicates the maximum severity recorded for each patient, rather than the severity recorded at admission. We find that, on average, the first viral load measurement for patients with mild disease was recorded earlier than those for either patients with moderate disease, or severe disease (p-values from a two-sided Wilcoxon test). b) Quantification of the first viral load measurement for each patient, stratified by severity classification. c) Quantification of the maximum viral load. As shown in the left-hand panel, we have many more data points for mild patients early after the onset of symptoms, which is why both the first and maximum recorded viral loads are higher on average for this group. Across these studies, 421 patients had more than one sample recorded: for 60.1% of these patients, the first sample had the highest recorded viral load.

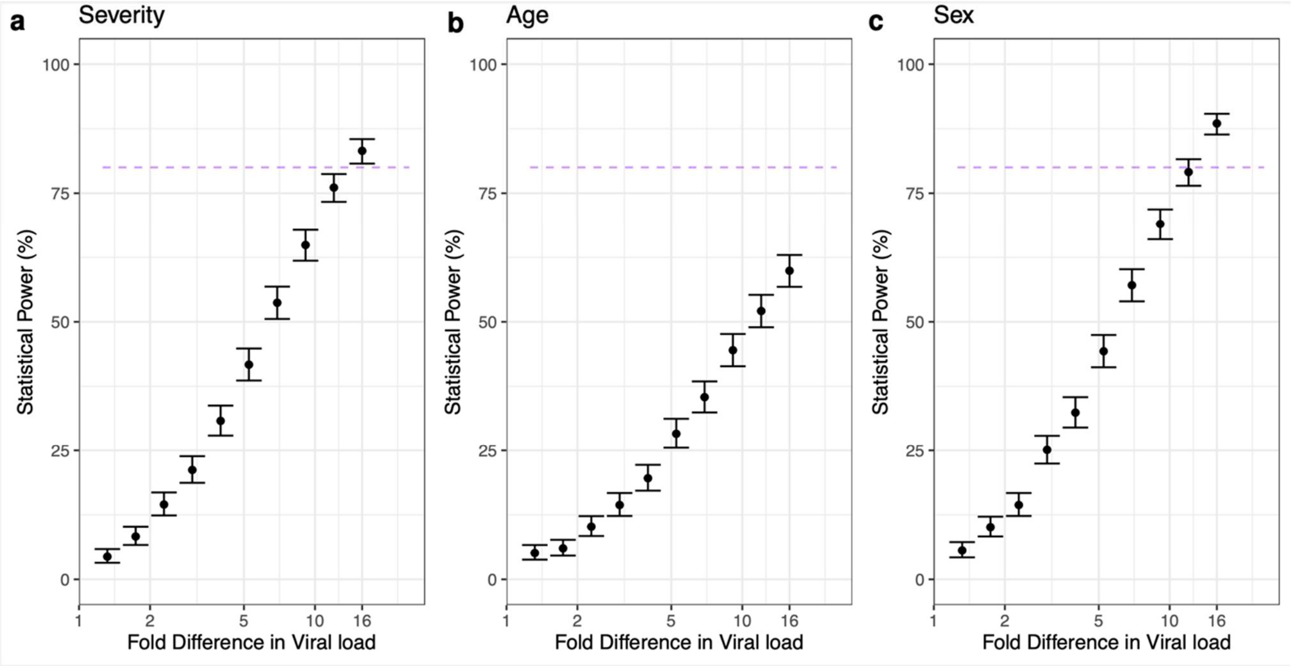


**Supplementary Figure 4:** Estimations of the statistical power in the regression analyses. We used simulation-based methods to estimate our power to measure an effect on the peak viral load due to severity of disease, age, or sex. We simulated datasets of the same size as the one used here for the regression analysis, containing the same level of variation between subjects and studies (Supplementary Table 3). In each set of simulations, the peak viral load was influenced by one of these 3 variables (panel (a): severity of disease; panel (b): age; panel (c): sex). In each case 1000 simulations were generated and the statistical power (black dots) calculated as the proportion of cases for which a significant effect (a fixed effect with a p value < 0.05 was deemed to show significance). The error bars show the 95% confidence intervals for each proportion calculated. In each panel, the purple, dashed line indicates 80% power. These plots suggest that we were underpowered to detect relatively small differences in viral load due to age, sex, or disease severity.

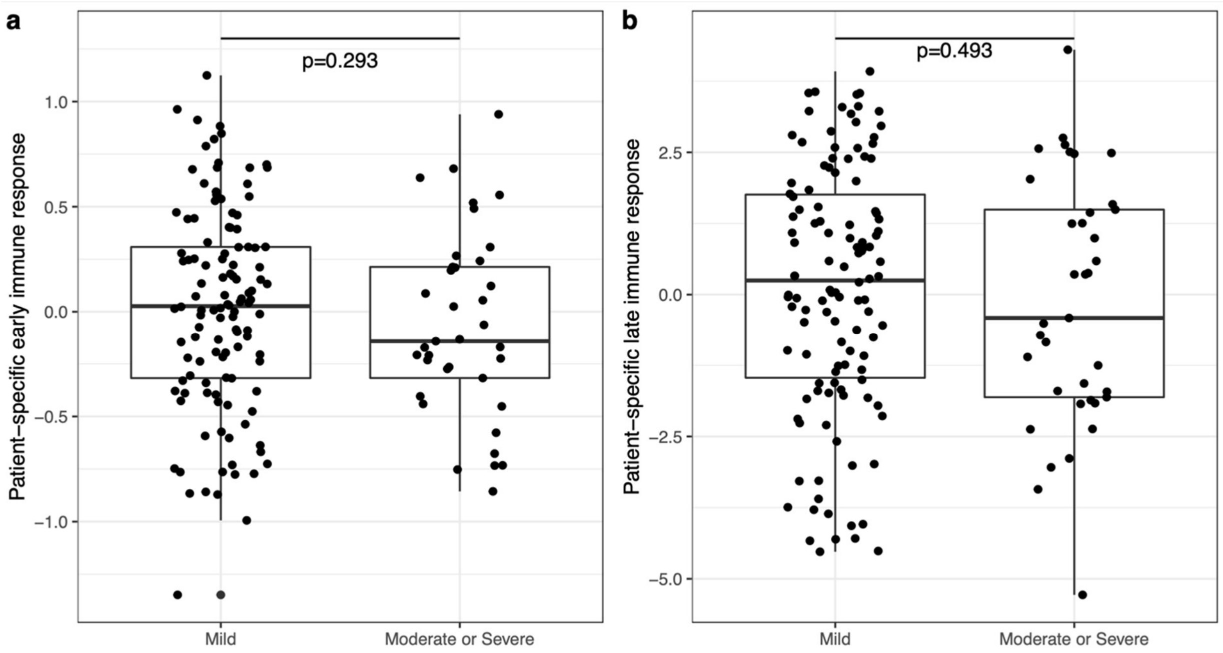


**Supplementary Figure 5**: Relationship between patient-specific parameters governing the immune response in the mechanistic model and disease severity. After adjusting for study (through the study-specific random effect) no significant differences were found between either the patient-specific early (panel a) or late (panel b) immune response parameters in the severity groups. This is consistent with the findings from the regression modelling. In this plot we show the posterior mean value of the subject-specific parameters.

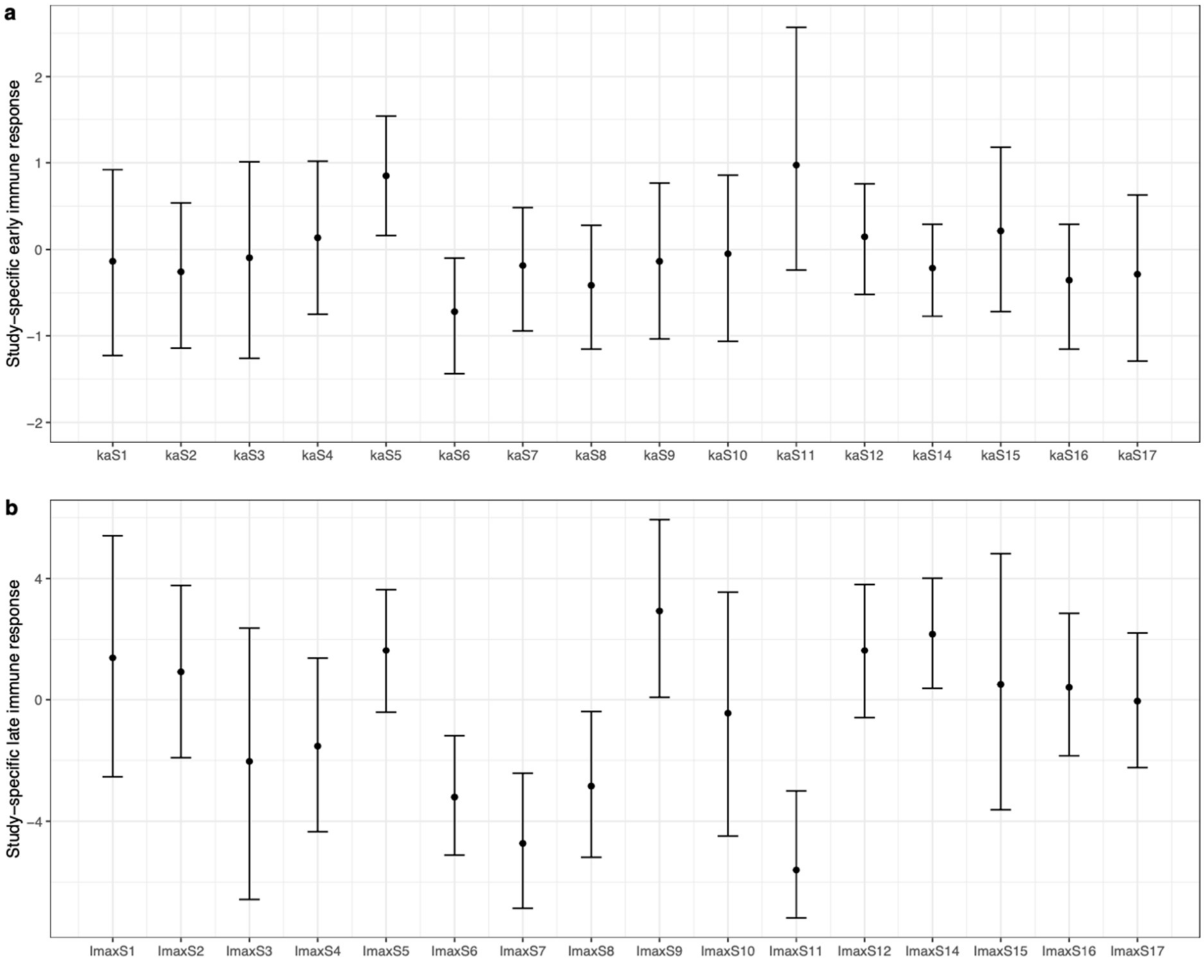


**Supplementary Figure 6**: Posterior means and 95% credible intervals for the study-specific offsets in the mechanistic model. Panel (a): the parameter values for the early immune response, which influences the peak viral load. Panel (b): the parameter values for the late immune response, which governs the rate at which the viral load is cleared.

**Supplementary Figure 7** *(shown over the following 7 pages)*: Output from the mechanistic model alongside the data, for all 155 patients considered. In the heading of each panel, the first number indicates the study (studies numbered as in Table 1). The second number identifies the patient. The coloured points indicate the data: for illustrative purposes, samples that were negative for virus are set to 1 viral copy per ml. The black lines indicate the model fit for each patient (calculated from the posterior mean). The shaded area shows the 95% credible intervals for the model fit.

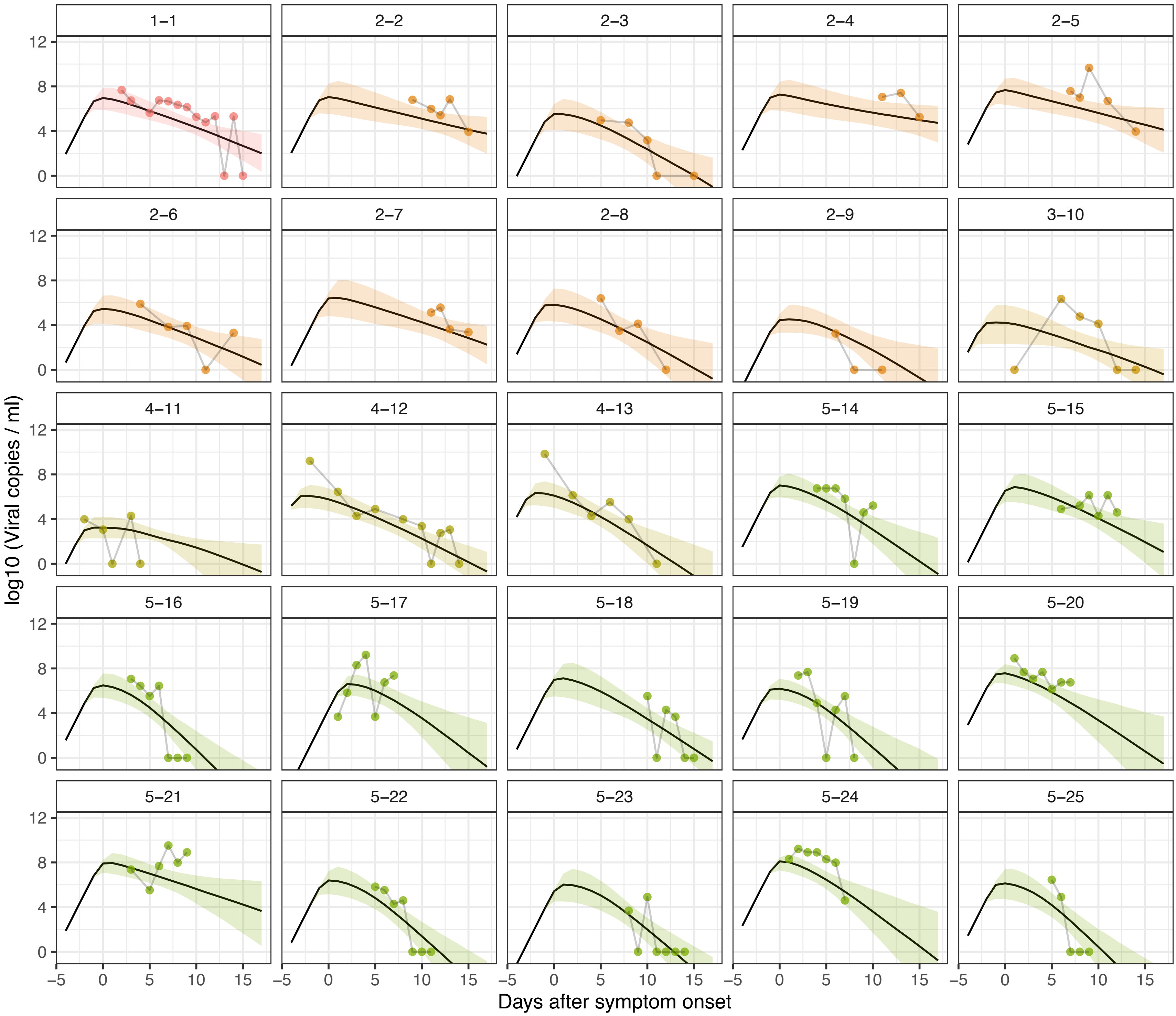

1–1
2–2
2–3
2–4
2–5
2–6
2–7
2–8
2–9
3–10
4–11
4–12
4–13
5–14
5–15
5–16
5–17
5–18
5–19
5–20
5–21
5–22
5–23
5–24
5–25
log10 (Viral copies / ml)
0
4
8
12
−5
0
5
10
15
Days after symptom onset

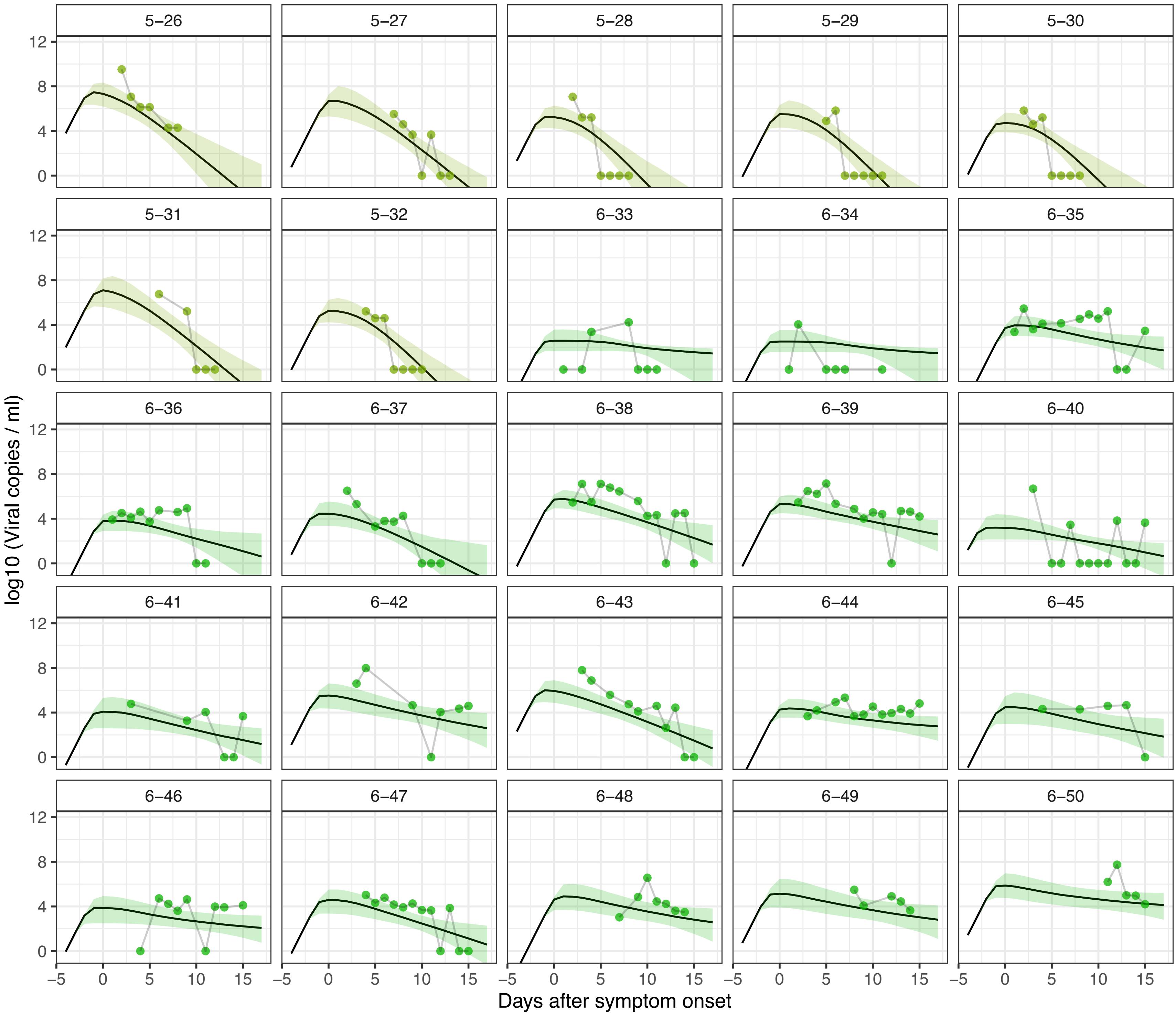
5−26
5−27
5−28
5−29
5−30
5−31
5−32
6−33
6−34
6−35
6−36
6−37
6−38
6−39
6−40
6−41
6−42
6−43
6−44
6−45
6−46
6−47
6−48
6−49
6−50
log10 (Viral copies / ml)
Days after symptom onset
0
4
8
12
−5
0
5
10
15

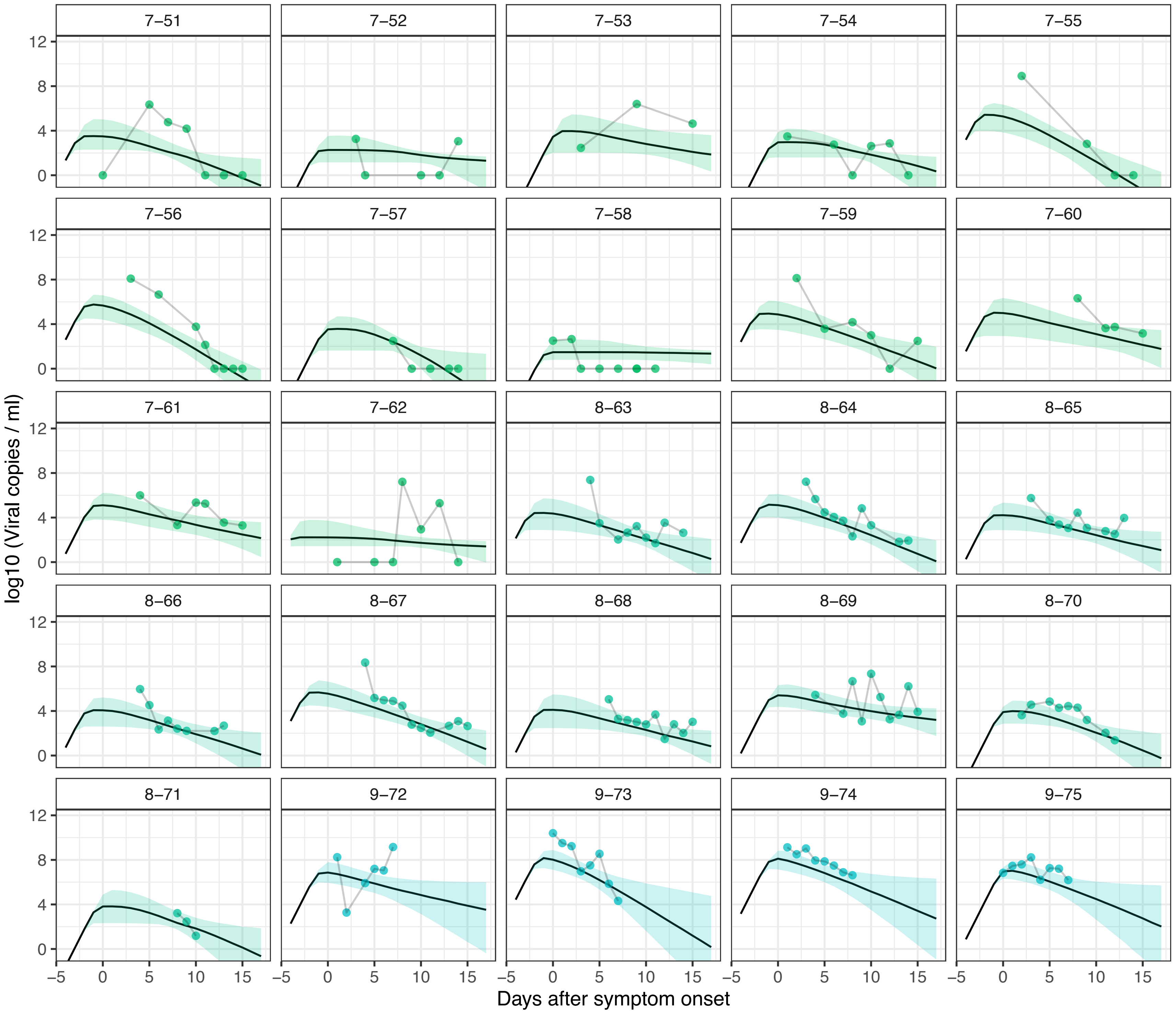
7−51
7−52
7−53
7−54
7−55
7−56
7−57
7−58
7−59
7−60
7−61
7−62
8−63
8−64
8−65
8−66
8−67
8−68
8−69
8−70
8−71
9−72
9−73
9−74
9−75
log10 (Viral copies / ml)
0
4
8
12
−5
0
5
10
15
Days after symptom onset

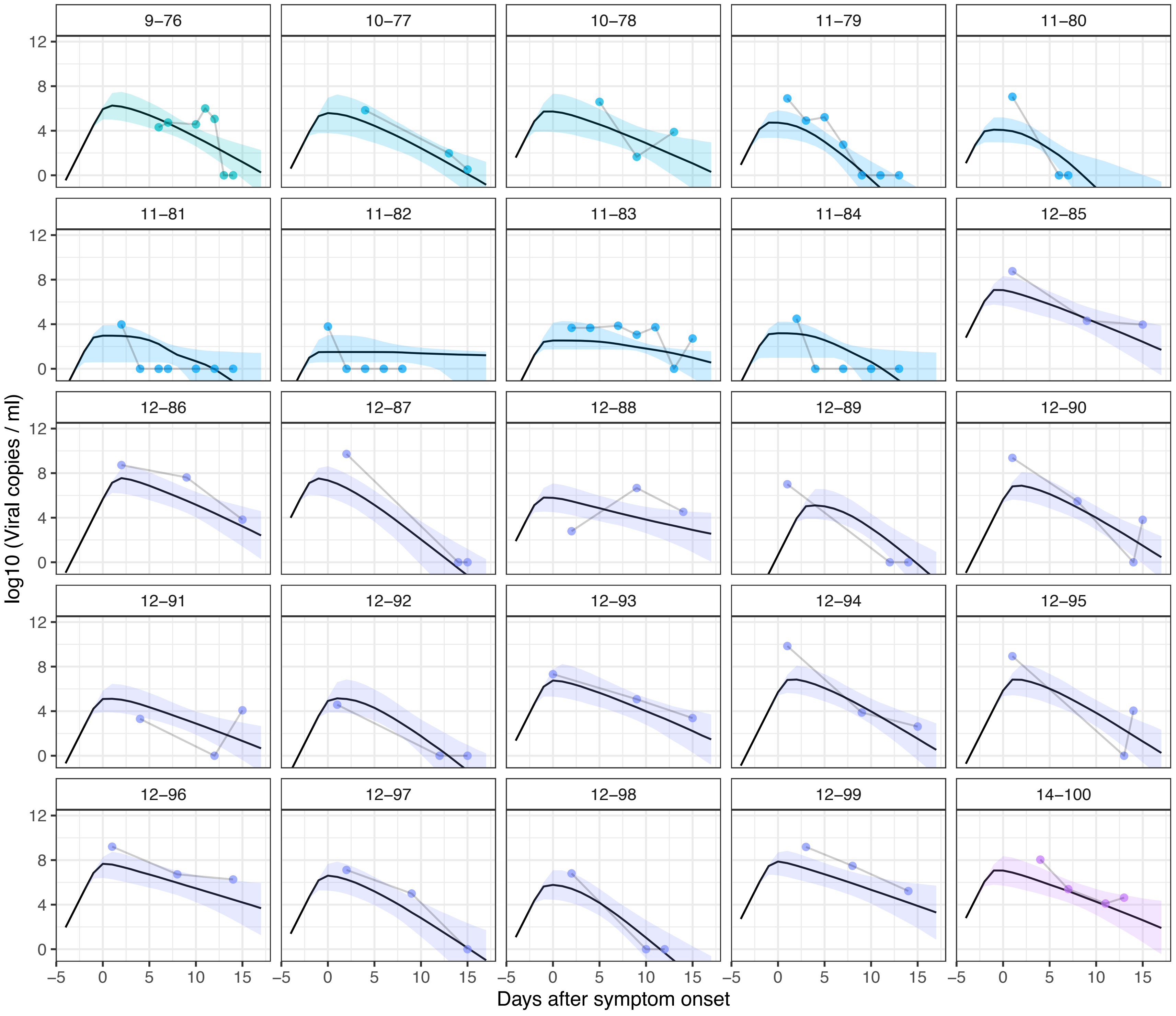

9–76
10–77
10–78
11–79
11–80
11–81
11–82
11–83
11–84
12–85
12–86
12–87
12–88
12–89
12–90
12–91
12–92
12–93
12–94
12–95
12–96
12–97
12–98
12–99
14–100
log10 (Viral copies / ml)
Days after symptom onset

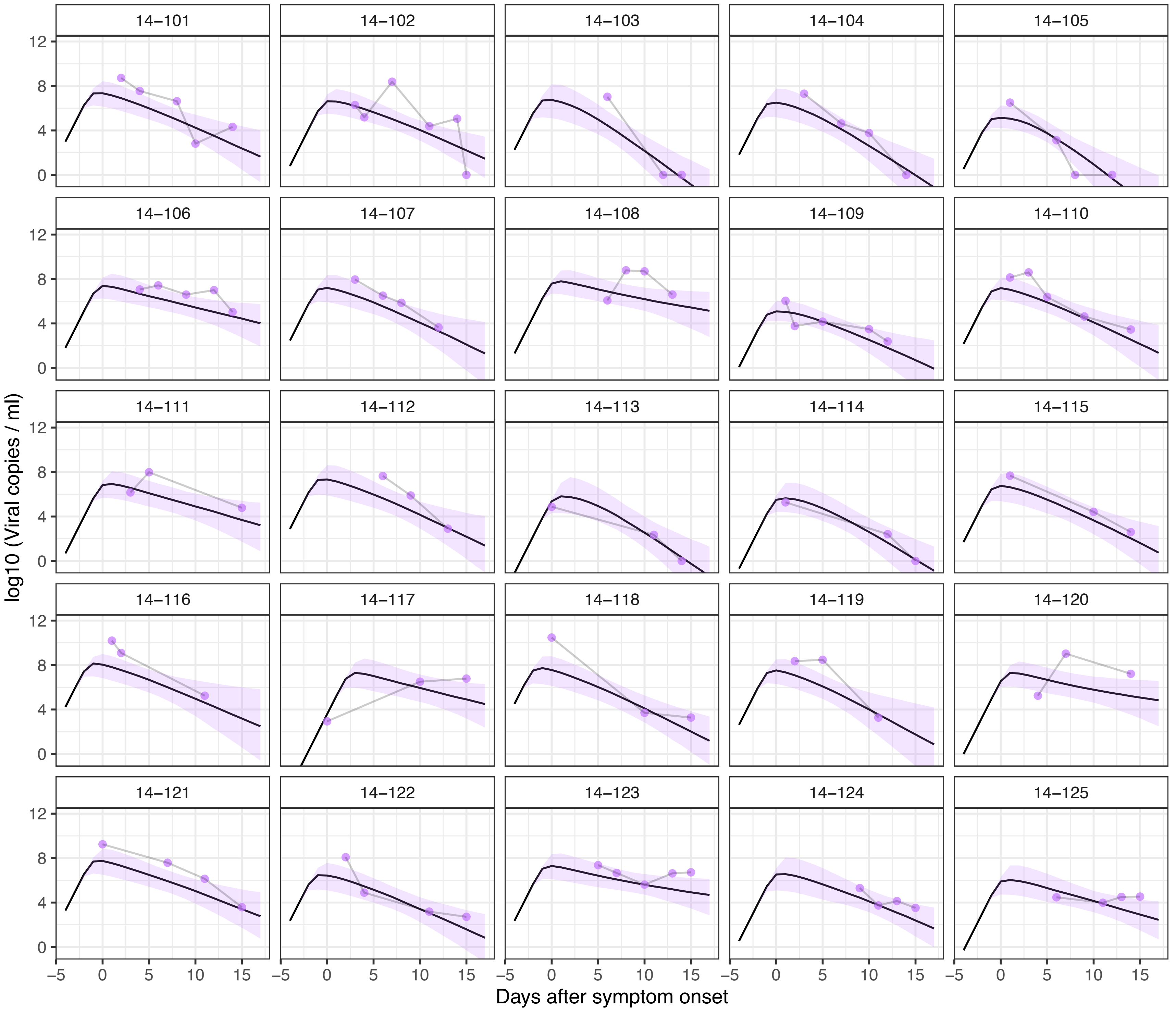
14−101
14−102
14−103
14−104
14−105
14−106
14−107
14−108
14−109
14−110
14−111
14−112
14−113
14−114
14−115
14−116
14−117
14−118
14−119
14−120
14−121
14−122
14−123
14−124
14−125
log10 (Viral copies / ml)
Days after symptom onset
0
4
8
12
−5
0
5
10
15

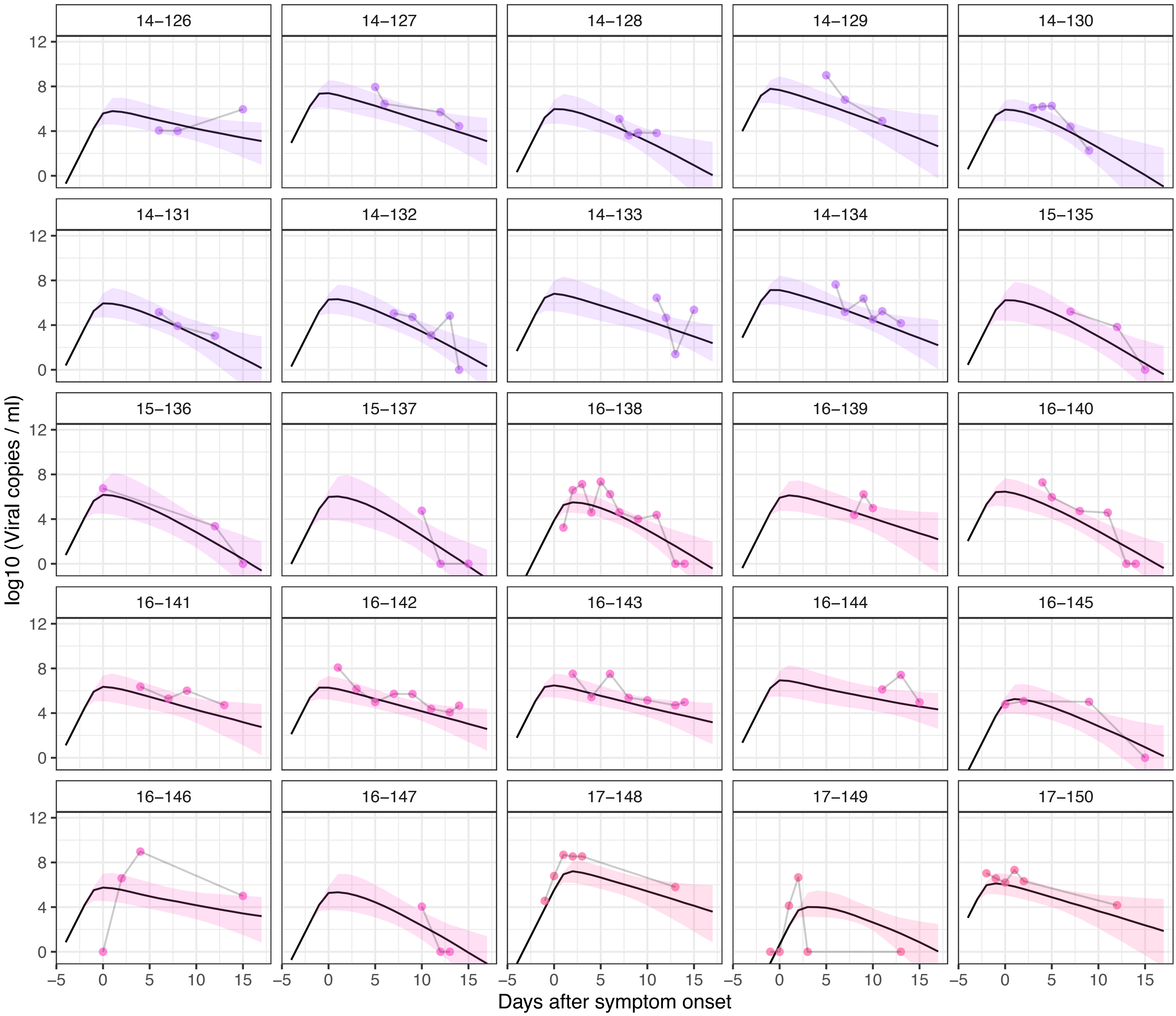
14–126
14–127
14–128
14–129
14–130
14–131
14–132
14–133
14–134
15–135
15–136
15–137
16–138
16–139
16–140
16–141
16–142
16–143
16–144
16–145
16–146
16–147
17–148
17–149
17–150
log10 (Viral copies / ml)
Days after symptom onset
0
4
8
12
–5
0
5
10
15

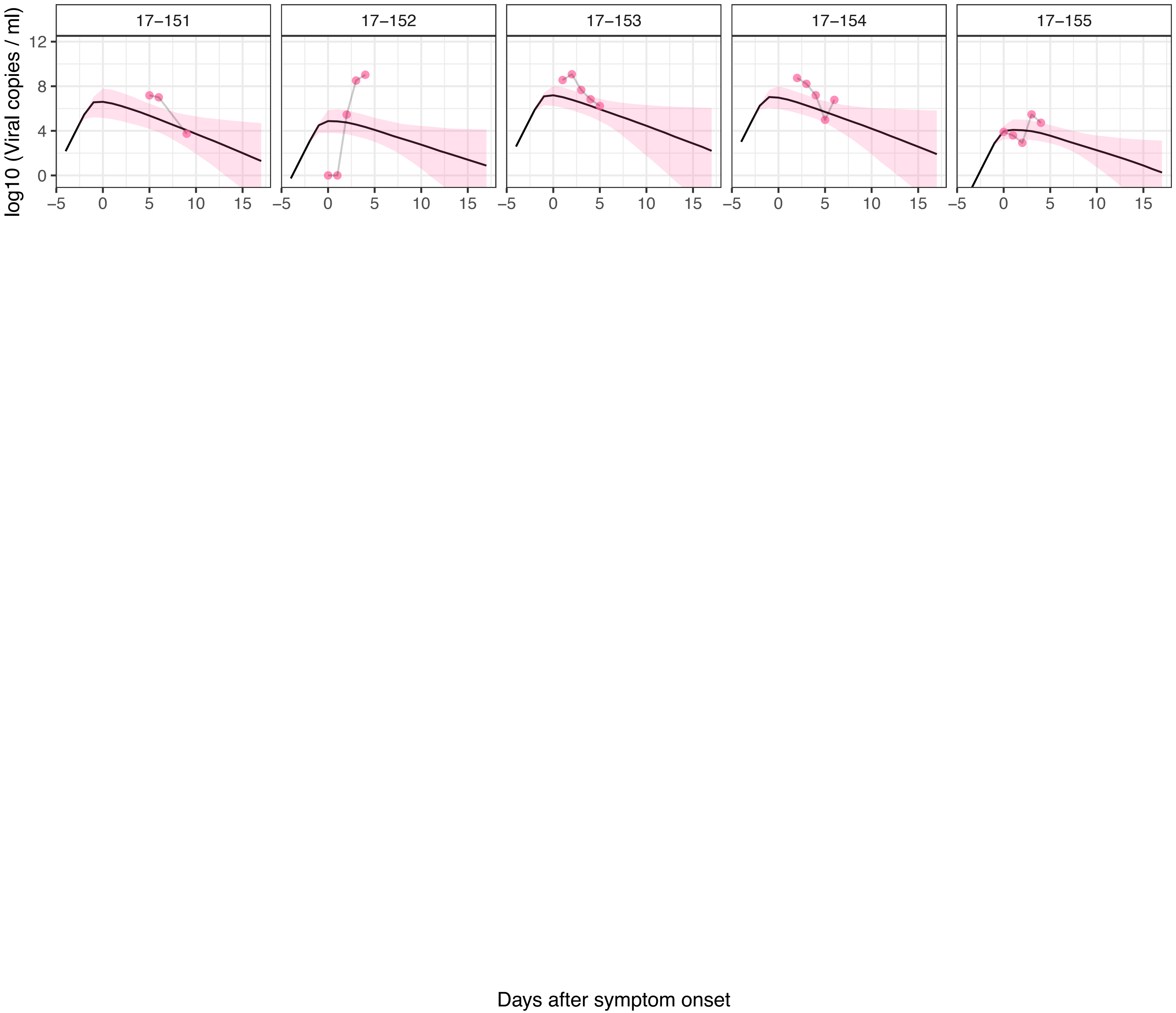
17−151
17−152
17−153
17−154
17−155
log10 (Viral copies / ml)
Days after symptom onset
